# The tryptophan catabolite or kynurenine pathway in autism spectrum disorder; a systematic review and meta-analysis

**DOI:** 10.1101/2023.05.07.23289630

**Authors:** Abbas F. Almulla, Yanin Thipakorn, Chavit Tunvirachaisakul, Michael Maes

**Author notes:** **Corresponding author:** Prof. Dr Michael Maes, M.D., Ph.D., Department of Psychiatry, Faculty of Medicine, Chulalongkorn University Bangkok, 10330, Thailand. **E-mail addresses:**.

## Abstract

**Background:** Autism spectrum disorder (ASD) is a neurodevelopment disorder characterized by impaired social communication and interaction, as well as rigid and unchanging interests and behaviors. In ASD, studies show activated immune-inflammatory and nitro-oxidative pathways which are accompanied by depletion of plasma tryptophan (TRP), increased competing amino acids (CAAs) and activation of the TRP catabolite (TRYCAT) pathway.

**Objectives:** This study aims to systematically review and meta-analyze data on peripheral TRP, CAAs, TRYCAT pathway activity, and individual TRYCATs, including kynurenine (KYN) and kynurenic acid (KA) levels, in blood and urine of ASD patients.

**Methods:** After searching PubMed, Google Scholar, and SciFinder extensively, a total of 25 full-text papers were included in the analysis, with a total of 6653 participants (3,557 people with ASD and 30,96 healthy controls).

**Results:** Blood TRP and the TRP/CAAs ratio were not significantly different between ASD patients and controls (standardized mean difference, SMD= −0.227, 95% confidence interval, CI: −0.540; 0.085 and SMD= 0.158, 95%CI: −0.042; 0.359) respectively. The KYN/TRP ratio showed no significant difference between ASD and controls (SMD= 0.001, 95%CI: −0.169; 0.171). Blood KYN and KA levels were not significantly changed in ASD. Moreover, there were no significant differences in urine TRP, KYN and KA levels between ASD and controls. We could not establish increases in neurotoxic TRYCATs in ASD.

**Conclusions:** Our study demonstrates that there are no abnormalities in peripheral blood TRP metabolism, IDO activity, and TRYCAT production in ASD. Reduced TRP availability and elevated neurotoxic TRYCAT levels are not substantial contributors to ASD’s pathophysiology.

## Introduction

Autism spectrum disorder (ASD) refers to a group of severe neurodevelopmental diseases known to cause a lifelong disability reflected by recurrent patterns of behavior and early difficulties with interpersonal and communicative skills (Wing and Gould 1979, American Psychiatric Association 2013). A recent meta-analysis reveals a global prevalence of 95% per 10000 individuals (Wang, Ma et al. 2022), indicating a dramatic rise in incidence since the 1960s and 1970s, when it was between 1% and 5% per 2,500 (Fombonne 2018, Wang, Ma et al. 2022). As the prevalence of ASD rises, families of ASD patients experience a growing financial and emotional burden (Ou, Shi et al. 2015, Picardi, Gigantesco et al. 2018).

Several hypotheses were proposed to explain the etiopathogenesis of ASD. (Yenkoyan, Grigoryan et al. 2017). The neuroimmune-inflammatory hypothesis of ASD has received a great deal of attention, as numerous researchers have pointed to an immune system imbalance in ASD patients (Warren, Margaretten et al. 1986, Croonenberghs, Bosmans et al. 2002, Ormstad, Bryn et al. 2018). Numerous studies and meta-analyses demonstrate an activated immune response system (IRS) in ASD with increased production of macrophage and T helper (Th)1, Th2, and Th17 cytokines, such as tumor necrosis factor-(TNF)-α, migration inhibitory factor (MIF), interleukin (IL)-1, IL-6, IL-8, IL-7, IL-12p70, CCL11, and MCP-1, IL-1RA, and interferon (IFN)-γ (Croonenberghs, Bosmans et al. 2002, Masi, Quintana et al. 2015, Xu, Li et al. 2015, Zhao, Zhang et al. 2021, Nie, Han et al. 2023). Furthermore, some meta-analyses found that children with ASD have significantly elevated C-reactive protein (CRP) (Nadeem, Hussain et al. 2020, Yin, Wang et al. 2020, Gardner, Lee et al. 2021) along with decreased α-2-macroglobulin, ceruloplasmin and transferrin (Chauhan, Chauhan et al. 2004, Hergüner, Keleşoğlu et al. 2012, Gardner, Lee et al. 2021) and increased cortisol (Corbett, Schupp et al. 2010, Spratt, Nicholas et al. 2012). Additionally, it has been reported that in ASD, the activity of T regulatory (Treg) cells was significantly reduced together with lowered concentrations of IL-1RA and IL-10, two immunoregulatory cytokines which are essential components of the compensatory immunoregulatory response system (CIRS) (Saghazadeh, Ataeinia et al. 2019, Ashwood 2023, Nie, Han et al. 2023).

Increased oxidative and nitrosative stress (O&NS) is invariably associated with an IRS/CIRS ratio imbalance and, accordingly, many studies show enhanced O&NS pathways in ASD (Chauhan, Gu et al. 2011, Bjørklund, Meguid et al. 2020, Manivasagam, Arunadevi et al. 2020, Nadeem, Ahmad et al. 2020, Pangrazzi, Balasco et al. 2020, Tripathi, Kartawy et al. 2020, Liu, Lin et al. 2022). Activated IRS and O&NS pathways result in several adverse outcomes, including induction of indoleamine 2,3-dioxygenase enzyme (IDO) with subsequent peripheral and central depletion of tryptophan (TRP) and the generation of neuroactive TRP catabolites (TRYCATs) including kynurenine (KYN), kynurenic acid (KA), 3-hydroxykynurenine (3HK), anthranilic acid (AA), quinolinic acid (QA), xanthurenic acid (XA), and picolinic acid (PA) (see **Figure 1**) (Maes, Leonard et al. 2011, Almulla and Maes 2022). Since TRP is a precursor of 5-HT and melatonin, aberrant TRP degradation may result in depletion of serotonin and melatonin (Maes, Leonard et al. 2011), which are both implicated in the pathophysiology of ASD (Croonenberghs, Verkerk et al. 2005, Croonenberghs, Wauters et al. 2007, Gabriele, Sacco et al. 2014). On the other hand, depleted levels of plasma TRP protect against hyperinflammation by downregulating the IRS, reducing immune cell proliferation, and protecting against microbial invasion (Maes, Leonard et al. 2011).

**Figure 1:**
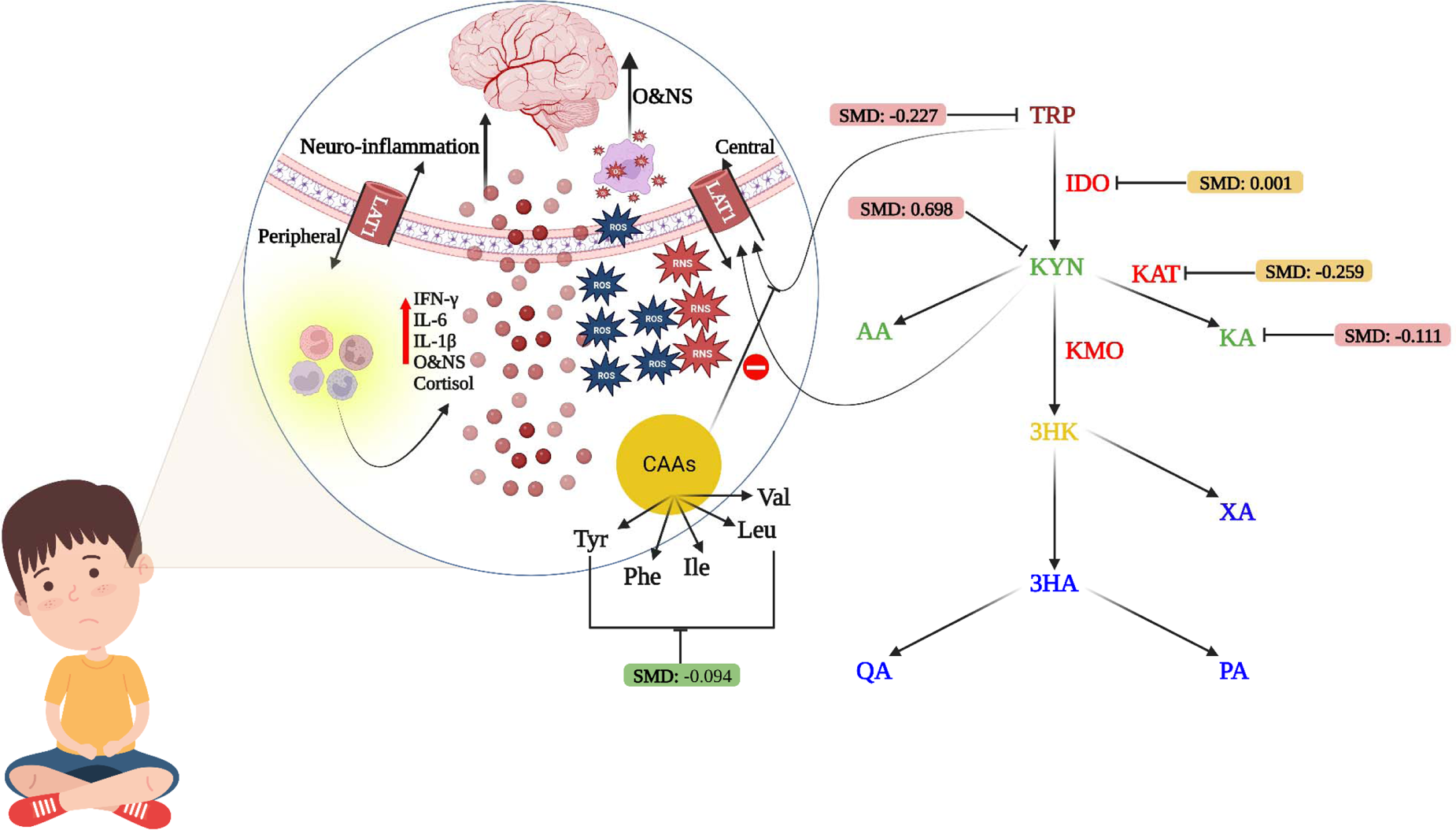
Summery of tryptophan (TRP), competing amino acids (CAAs), tryptophan catabolites (TRYCATs) results in autism spectrum disorder. CAAs: Competing amino acids, Ile: Isoleucine, Leu: Leucine, Val: Valine, Phe: Phenylalanine, Tyr: Tyrosine, O&NS: Oxidative and nitrosative stress, LAT1: Large neutral amino acid transporter 1, IFN: Interferon, IL: Interleukin, ROS: Reactive Oxygen Species, RNS: Reactive Nitrogen species. TRP: Tryptophan, KYN: Kynurenine, KA: Kynurenic acid, 3-HK: 3-Hydroxy KYN, QA: Quinolinic acid, XA: Xanthurenic acid, PA: Picolinic acid.

Some studies reported significantly diminished peripheral TRP levels in ASD patients (Croonenberghs, Delmeire et al. 2000, Adams, Audhya et al. 2011, Tu, Chen et al. 2012, Naushad, Jain et al. 2013, Bugajska, Berska et al. 2017). It should be noted that the delivery of TRP to the brain depends partially on the peripheral levels of competing amino acids (CAAs) and the ratio of TRP/CAA, whereby increased levels of CAAs, including leucine, isoleucine, valine, tyrosine and phenylalanine, may diminish the availability of TRP to the brain (Almulla, Vasupanrajit et al. 2022). A previous study showed a significantly decreased TRP/CAA ratio in ASD (D’Eufemia, Finocchiaro et al. 1995). Other studies, however, showed that TRP levels in the peripheral blood of ASD patients were comparable to those of healthy controls (Bryn, Verkerk et al. 2017, Ormstad, Bryn et al. 2018, Bilgiç, Abuşoğlu et al. 2022). Case-control studies did not show a significant induction of the TRYCAT pathway in ASD as evaluated by the KYN/TRP ratio, an indicator of IDO (Bryn, Verkerk et al. 2017, Ormstad, Bryn et al. 2018, Bilgiç, Abuşoğlu et al. 2022). Regarding TRP in urine, some studies asserted an increased TRP urinary excretion in ASD (Noto, Fanos et al. 2014, Liang, Xiao et al. 2020), whereas other studies refuted these claims (Kałuzna-Czaplinska, Michalska et al. 2010, Li, Shen et al. 2018).

Intriguingly, researchers have discovered that several TRYCATs are considerably altered in ASD, including elevated serum QA and KA levels (Lim, Essa et al. 2016, Bilgiç, Abuşoğlu et al. 2022), whereas other studies reported significantly lower QA and KA levels in ASD (Bryn, Verkerk et al. 2017, Ormstad, Bryn et al. 2018, Carpita, Nardi et al. 2022).

No previous systematic review and meta-analysis examined peripheral TRP, TRP/CAA ratio, TRYCATs and activity of the IDO enzyme in ASD patients. Hence, the aim of the present meta-analysis is to investigate peripheral assessments of TRP, TRP/CAA, the KYN/TRP ratio and TRYCATs in ASD versus controls.

## Materials and Methods

In the present meta-analysis, we compare ASD patients and healthy controls in terms of TRP, TRP/CAAs, TRYCATs, TRYCAT ratios including KYN/TRP and KA/KYN ratios, and a composite score of neurotoxic TRYCATs (KYN + 3HK + XA + QA + PA) in serum/plasma and urine. The guidelines established by the Preferred Reporting Items for Systematic Reviews and Meta-Analyses (PRISMA) 2020 (Page, McKenzie et al. 2021), Cochrane Handbook for Systematic Reviews and Interventions (Higgins, Thomas et al. 2019), and the Meta-Analyses of Observational Studies in Epidemiology (MOOSE) were adhered to while carrying out the present meta-analysis.

### Search Strategy

The databases PubMed/MEDLINE, Google Scholar, and SciFinder were searched from March 10^th^, 2023 to the end of April 2023. Table 1 of the electronic supplementary file (ESF) demonstrates the keywords and mesh terms used to collect papers concerning TRP, CAAs and TRYCATs in ASD. However, we double-checked the reference lists of all qualifying articles and all previous meta-analyses to guarantee that no relevant research was missed.

**Table 1.** The outcomes and number of patients with autism spectrum disorder (ASD) and healthy control along with the side of standardized mean difference (SMD) and the 95% confidence intervals with respect to zero SMD.

| Outcome profiles | n<br>studies | Side of 95% confidence intervals |  |  |  | Patient<br>Cases | Control<br>Cases | Total<br>number of<br>participants |
| --- | --- | --- | --- | --- | --- | --- | --- | --- |
|  |  | < 0 | Overlap 0<br>and SMD<br>< 0 | Overlap 0<br>and SMD<br>> 0 | > 0 |  |  |  |
| Serum/Plasma |  |  |  |  |  |  |  |  |
| TRP/CAAs | 16 | 2 | 4 | 4 | 6 | 2317 | 2254 | 4571 |
| TRP | 16 | 6 | 3 | 6 | 1 | 834 | 855 | 1689 |
| CAAs | 7 | 1 | 5 | 0 | 1 | 1483 | 1399 | 2882 |
| KYN/TRP | 3 | 0 | 2 | 1 | 0 | 400 | 200 | 600 |
| KA/KYN | 5 | 2 | 2 | 1 | 0 | 478 | 272 | 750 |
| KYN | 4 | 0 | 0 | 3 | 1 | 215 | 112 | 327 |
| KA | 5 | 3 | 1 | 1 | 0 | 263 | 160 | 423 |
| (KYN+ 3-HK + QA+ XA+ PA) | 4 | 0 | 2 | 2 | 0 | 510 | 266 | 776 |
| Urine |  |  |  |  |  |  |  |  |
| TRP | 4 | 2 | 0 | 0 | 2 | 80 | 91 | 171 |
| KYN | 2 | 2 | 0 | 0 | 0 | 44 | 44 | 88 |
| KA | 2 | 1 | 1 | 0 | 0 | 44 | 44 | 88 |

SMD: Standard Mean Difference TRP: Tryptophan, KYN: Kynurenine, KA: Kynurenic acid, CAAs: Competing amino acids (Leucine, Isoleucine, Valine, Phenylalanine, Tyrosine), 3-HK: 3-Hydroxy KYN, QA: Quinolinic acid, XA: Xanthurenic acid, PA: Picolinic acid.

### Eligibility Criteria

This meta-analysis included only articles that met two essential criteria: publication in peer-reviewed journals and English language writing. We searched for grey literature, which included articles written in Thai, French, Spanish, German, Italian, and Russian, even though all the relevant publications were written in English and peer reviewed. We included case-control and cohort studies that measured TRP, CAAs, and TRYCAT levels in serum, plasma, or urine samples and examined autistic people who were diagnosed using the criteria established by the Diagnostic and Statistical Manual of Mental Disorders (DSM) or the International Classification of Diseases (ICD). Longitudinal studies could be included if serum/plasma or urinary TRP, CAAs and TRYCATs were assessed in baseline conditions. We excluded: a) systematic reviews and meta-analyses, as well as animal, translational and genetic-based studies, b) studies that lacked a control group or that presented replicated findings, c) studies that measured the biomarkers in saliva, whole blood, platelet-rich plasma, and blood cells, and d) articles that did not provide mean, standard deviation (SD) or standard error (SE) of the biomarkers. However, we requested that the authors supply us with the mean (SD/SE) values if they were absent from their publication. In the absence of author-supplied data, we calculated means and SDs from graphical forms using the Web Plot Digitizer (https://automeris.io/WebPlotDigitizer/) or approximated means and SDs from medians using the Wan et al. (2014) method (Wan, Wang et al. 2014).

### Primary and secondary outcomes

The primary outcome of the present meta-analysis was the evaluation of serum/plasma levels of TRP and KYN as well as the KYN/TRP ratio as an indicator of IDO enzyme activity, as shown in Table 1. We also obtained secondary outcomes by estimating KAT enzyme indices (KA/KYN ratio and solitary levels of KA) and a neurotoxic TRYCAT composite score (KYN + 3HK+3HA + QA + XA + PA). Urinary levels of TRP and KYN were also assessed in the present study.

### Screening and data extraction

The first two authors (AA and YT) reviewed the titles and abstracts of the relevant articles to evaluate whether they met the inclusion criteria for this meta-analysis. Full-text papers that met our inclusion criteria were therefore downloaded, whereas those that did not were ignored. The mean (SD) TRYCAT data, and other pertinent clinical data from the included studies were entered in a standard Excel spreadsheet. The prespecified Excel file included author names, study dates, TRP and TRYCAT names, patient and healthy controls group sample sizes, media type (plasma, serum, and urine), psychiatric assessment scales, and demographic information such as the mean (SD) age, gender, and location of the study (latitude). YT and AA double-checked the spreadsheet and contacted MM to check for any discrepancies.

After being modified by the corresponding author (MM) for usage with the research under our investigation, the Immunological Confounder Scale (ICS) was utilized to evaluate the study’s methodological quality (Andrés-Rodríguez, Borràs et al. 2020). ESF, Table 2 describes two scales that assessed the quality of the research using the quality and red points scales. We frequently utilized these rating scales to assess the methodological quality of articles examining the TRP and TRYCATs in affective disorders (Almulla, Thipakorn et al. 2022, Almulla, Thipakorn et al. 2022), schizophrenia (Almulla, Vasupanrajit et al. 2022), and Alzheimer’s disease (Almulla, Supasitthumrong et al. 2022). The quality scale ranges from 0 to 10, with the best quality attained as the score approaches 10. In this scale, sample size, controlling for confounders, and duration of sampling were primarily considered. In contrast, the primary objective of the red point scale is to predict the likelihood of bias in the results of biomarker assays and study design by evaluating the degree to which critical confounders were controlled for. The level of control is at its highest when the overall score is zero; whereas when it is twenty-six, the confounding variables are not taken into consideration.

**Table 2.** Results of meta-analysis performed on several outcome (TRYCATs) variables with combined serum/plasma and separate urine media

| Outcome feature sets | n | Groups | SMD | 95% CI | z | p | Q | df | p | I<sup>2</sup> (%) | $\tau^2$ | T |
| --- | --- | --- | --- | --- | --- | --- | --- | --- | --- | --- | --- | --- |
| **Serum/Plasma** |  |  |  |  |  |  |  |  |  |  |  |  |
| TRP/CAAs | 16 | Overall | 0.158 | -0.042; 0.359 | 1.549 | 0.121 | 123.727 | 15 | 0.000 | 87.877 | 0.131 | 0.361 |
| TRP | 16 | Overall | -0.227 | -0.540; 0.085 | -1.425 | 0.154 | 119.627 | 15 | 0.000 | 87.461 | 0.336 | 0.580 |
| CAAs | 7 | Overall | -0.094 | -0.272; 0.085 | -1.029 | 0.304 | 27.097 | 6 | 0.000 | 77.857 | 0.042 | 0.205 |
| KYN/TRP | 3 | Overall | 0.001 | -0.169; 0.171 | 0.014 | 0.989 | 0.360 | 2 | 0.835 | 0.000 | 0.000 | 0.000 |
| KA/KYN | 5 | Overall | -0.259 | -0.520; 0.001 | -1.949 | 0.051 | 10.553 | 4 | 0.032 | 62.095 | 0.053 | 0.229 |
| KYN | 4 | Overall | 0.698 | -0.061; 1.457 | 1.802 | 0.072 | 26.915 | 3 | 0.000 | 88.854 | 0.502 | 0.708 |
| KA | 5 | Overall | -0.111 | -0.585; 0.362 | -0.461 | 0.645 | 20.859 | 4 | 0.000 | 80.824 | 0.229 | 0.479 |
| (KYN+ 3-HK + QA+<br>XA+ PA) | 4 | Overall | -0.018 | -0.167; 0.131 | -0.241 | 0.809 | 1.970 | 3 | 0.579 | 0.000 | 0.000 | 0.000 |
| **Urine** |  |  |  |  |  |  |  |  |  |  |  |  |
| TRP | 4 | Overall | -0.201 | 2.457; 2.054 | -0.175 | 0.861 | 107.872 | 3 | 0.000 | 97.219 | 5.132 | 2.265 |
| KYN | 2 | Overall | -1.021 | -1.465; -0.577 | -4.504 | 0.000 | 0.055 | 1 | 0.814 | 0.000 | 0.000 | 0.000 |
| KA | 2 | Overall | -1.668 | -3.912; 0.576 | -1.457 | 0.145 | 18.720 | 1 | 0.000 | 94.658 | 2.482 | 1.575 |

SMD: Standard Mean Difference TRP: Tryptophan, KYN: Kynurenine, KA: Kynurenic acid, CAAs: Competing amino acids (Leucine, Isoleucine, Valine, Phenylalanine, Tyrosine), 3-HK: 3-Hydroxy KYN, QA: Quinolinic acid, XA: Xanthurenic acid, PA: Picolinic acid.

### Data analysis

At least two studies are required to conduct a meta-analysis on the TRP or TRYCAT data. The current meta-analysis assumed dependence to compare mean TRP/CAAs, KYN/TRP, and KA/KYN ratios, and neurotoxicity indicator values between individuals with ASD and healthy controls. The TRP/CAAs, KYN/TRP, and KA/KYN were calculated to assess the availability of TRP to the brain and the activity of the IDO and KAT enzymes (Almulla, Thipakorn et al. 2022, Almulla, Vasupanrajit et al. 2022). For the KYN/TRP analysis reflecting IDO activity, we set the effect sizes to be negative for decreased TRP and positive for increased KYN. Likewise, a meta-analysis was used to identify KAT enzyme activity, with a positive effect size for increased KA and a negative effect size for decreased KYN. In addition, the ratio of TRP/CAAs was determined by considering a positive effect size for TRP and a negative effect size for the CAAs.

Given the heterogeneity in participant characteristics between studies, we pooled effect values using a random-effects model with restricted maximum likelihood. A two-tailed p-value of less than 0.05 was deemed statistically significant, and the effect size was assigned as the SMD with 95% CI. SMD values of 0.80, 0.50, and 0.20 indicated a large, moderate, and small effect size, respectively (Cohen 2013). As in previous meta-analyses, heterogeneity was measured using tau-squared statistics; however, we also provide Q and I^2^ measures (Almulla, Supasitthumrong et al. 2022, Almulla, Thipakorn et al. 2023). Additionally, a meta-regression was used to identify the causes of heterogeneity. Subgroup analysis was performed to see if there were significant variations in TRP, CAAs, and TRYCATs concentrations between serum, plasma, and urine after selecting each of them as a unit of analysis. Using the leave-one-out sensitivity analysis, we checked the consistency of effect sizes and heterogeneity across studies.

In this meta-analysis, publication bias was examined using the fail-safe N method, continuity-corrected Kendall tau, and Egger’s regression intercept, with one-tailed p-values for the last two approaches. When Egger’s test showed an asymmetry, we imputed the missing studies and computed adjusted effect sizes using the trim-and-fill method (Duval and Tweedie 2000). Since funnel plots (study precision vs SMD) show both observed and imputed missing values, they were also used to locate small study effects. This meta-analysis was performed using the CMA V3 software and in accordance with the PRISMA criteria (ESF, Table 3).

**Table 3.** Results on publication bias.

| Outcome feature sets | Fail safe n | Z Kendall's $\tau$ | p | Egger's t test (df) | p | Missing studies (side) | After Adjusting |
| --- | --- | --- | --- | --- | --- | --- | --- |
| TRP/CAAs | 2.755 | 0.675 | 0.249 | 1.662(14) | 0.059 | 1 (Left) | 0.111 (-0.089; 0.313) |
| TRP | -4.575 | 0.045 | 0.482 | 0.990(14) | 0.169 | 0 | - |
| CAAs | -1.692 | 0.600 | 0.274 | 1.03 (5) | 0.175 | 2 (Right) | 0.002 (-0.176; 0.182) |
| KYN/TRP | -0.016 | 1.044 | 0.148 | 75.12(1) | 0.004 | 0 | - |
| KA/KYN | 3.138 | 2.204 | 0.013 | 2.222(3) | 0.056 | 2 (Right) | -0.126 (-0.381; 0.127) |
| KYN | 3.851 | 0.339 | 0.367 | 4.771(2) | 0.021 | 1 (Right) | 1.028 (0.120; 1.936) |
| KA | -1.153 | 0.000 | 0.500 | 0.265(3) | 0.404 | 1 (Left) | -0.311 (-0.875; 0.252) |
| (KYN+ 3-HK + QA+ XA+ PA) | -0.219 | 0.000 | 0.500 | 0.092(2) | 0.467 | 0 | - |
| **Urine** |  |  |  |  |  |  |  |
| TRP | -0.959 | 0.000 | 0.500 | 0.027(2) | 0.490 | 0 | - |

TRP: Tryptophan, KYN: Kynurenine, KA: Kynurenic acid, CAAs: Competing amino acids (Leucine, Isoleucine, Valine, Phenylalanine, Tyrosine), 3-HK: 3-Hydroxy KYN, QA: Quinolinic acid, XA: Xanthurenic acid, PA: Picolinic acid.

## Results

### Search results

ESF, Table 1 shows the keywords and Mesh terms that were employed to perform the search process that led to the identification of 50392 studies. The PRISMA flow diagram, depicted in **Figure 2**, provides a representation of the search outcomes by determining the total count of included and omitted studies. However, after additional screening to remove irrelevant studies, we were left with 473 articles. Only 30 research articles met the inclusion criteria for this systematic review; 444 were excluded because they did not meet the criteria. In addition, 4 research articles were disqualified for reasons listed in ESF, Table 4. As a result, 25 studies were included in this meta-analysis since they fulfilled the predetermined inclusion-exclusion criteria (Hoshino, Yamamoto et al. 1984, Hoshino, Yamamoto et al. 1986, D’Eufemia, Finocchiaro et al. 1995, Croonenberghs, Delmeire et al. 2000, Kałuzna-Czaplinska, Michalska et al. 2010, Adams, Audhya et al. 2011, Tu, Chen et al. 2012, Naushad, Jain et al. 2013, ElBaz, Zaki et al. 2014, Noto, Fanos et al. 2014, Gevi, Zolla et al. 2016, Lim, Essa et al. 2016, Bryn, Verkerk et al. 2017, Bugajska, Berska et al. 2017, Li, Shen et al. 2018, Ormstad, Bryn et al. 2018, Liang, Xiao et al. 2020, Olesova, Galba et al. 2020, Bilgiç, Abuşoğlu et al. 2022, Carpita, Nardi et al. 2022, Gagliano, Murgia et al. 2022, Kalejahi, Kheirouri et al. 2022, Raghavan, Anand et al. 2022, Timperio, Gevi et al. 2022, Chen, Chen et al. 2023).

**Figure 2:**
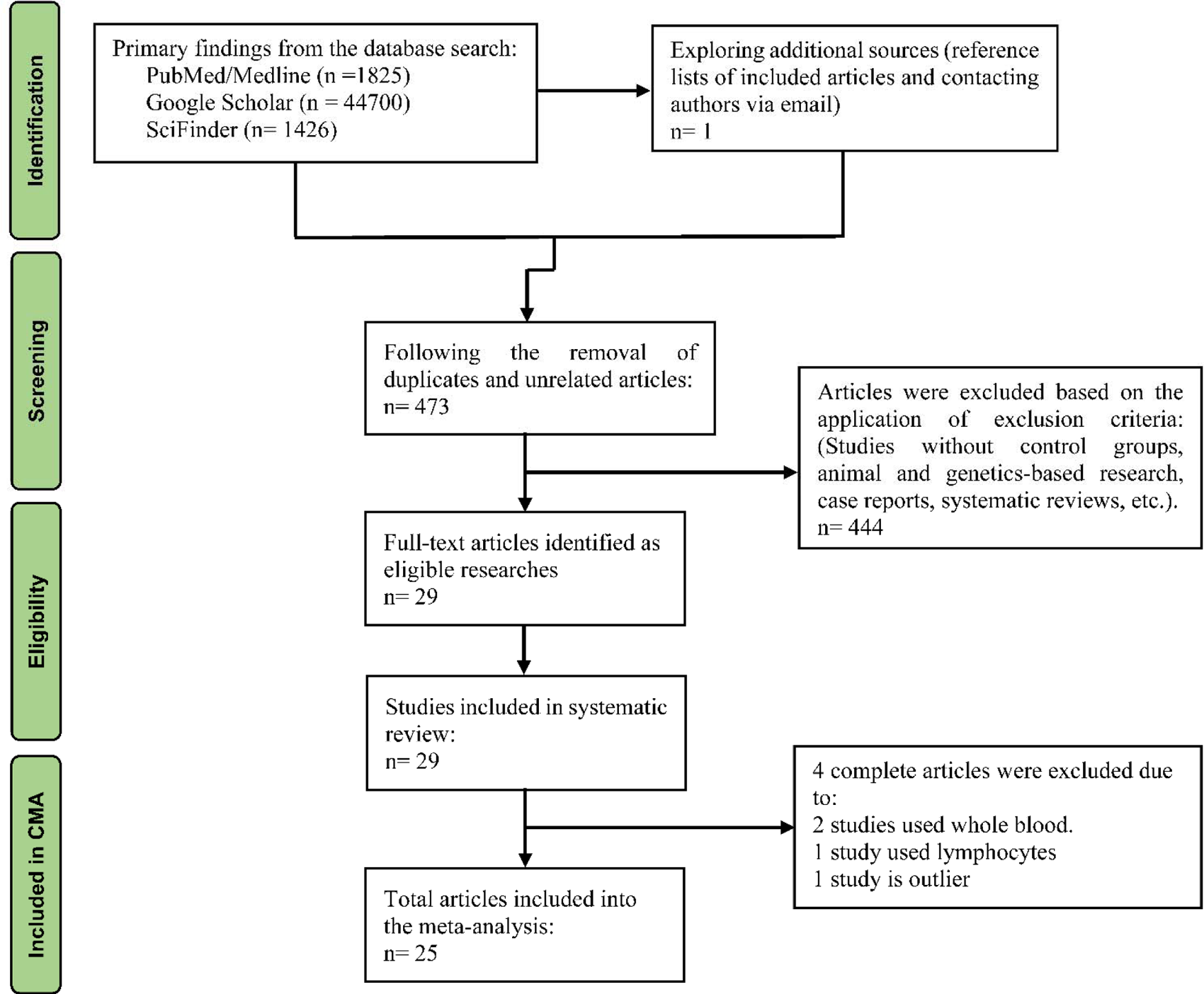
The PRISMA flow chart

In this meta-analysis, we involved 25 studies to compute the overall effect size (18 serum/plasma and 7 urine studies). There were 6653 participants in total, including 3096 healthy controls people and 3557 autistic patients. The ages of the participants ranged from 2.5 to 55.5 years. High-performance liquid chromatography was more frequently used than enzyme-linked immunosorbent assay (ELISA) and liquid chromatography with tandem mass spectrometry (LC/MS-MS) (see ESF, Table 5). Italy contributed the most studies (six), followed by China (four) and the United States (three), in diminishing order of contribution quantity. The median (min-max) values for quality and redpoint scores and shown in ESF, table 5.

### Primary outcome variables

#### TRP/CAA ratio, CAAs, and TRP in ASD

Sixteen studies examined the TRP/CAAs ratio as shown in **Table 1**. **Table 2** and **Figure 3** display that patients with ASD do not show any changes in the TRP/CAAs ratio as compared with healthy controls. However, there was some bias with 1 missing study to the left of the funnel plot and adjusting the effect size decreased the SMD (see **Table 3**). Table 1 and Table 2 and ESF, Figure 1 show no significant changes in CAA levels in ASD. We extracted the effect size of TRP from 16 studies.

**Figure 3.**
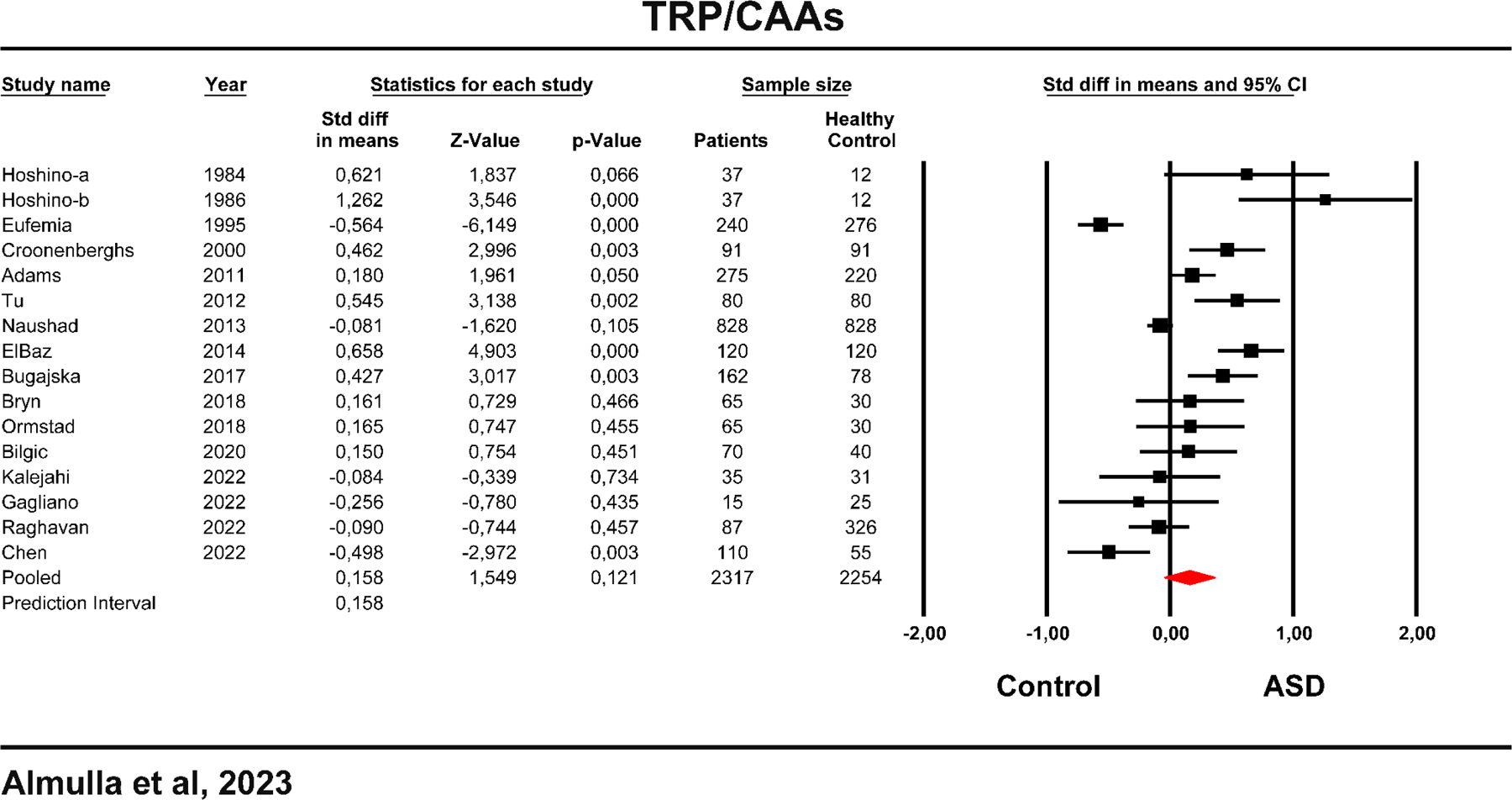
Forest plot of tryptophan (TRP)/competing amino acids (CAAs) in the patients with autism spectrum disorder (ASD) versus healthy controls.

Table 2 and **Figure 4** show that the TRP levels did not significantly differ between ASD and healthy controls. Publication bias was absent in TRP results. In addition, we examined urinary TRP levels. The results in Table 1 and Table 2 revealed no significant difference in urinary TRP between ASD and controls. Table 3 indicates no bias in urinary TRP results.

**Figure 4:**
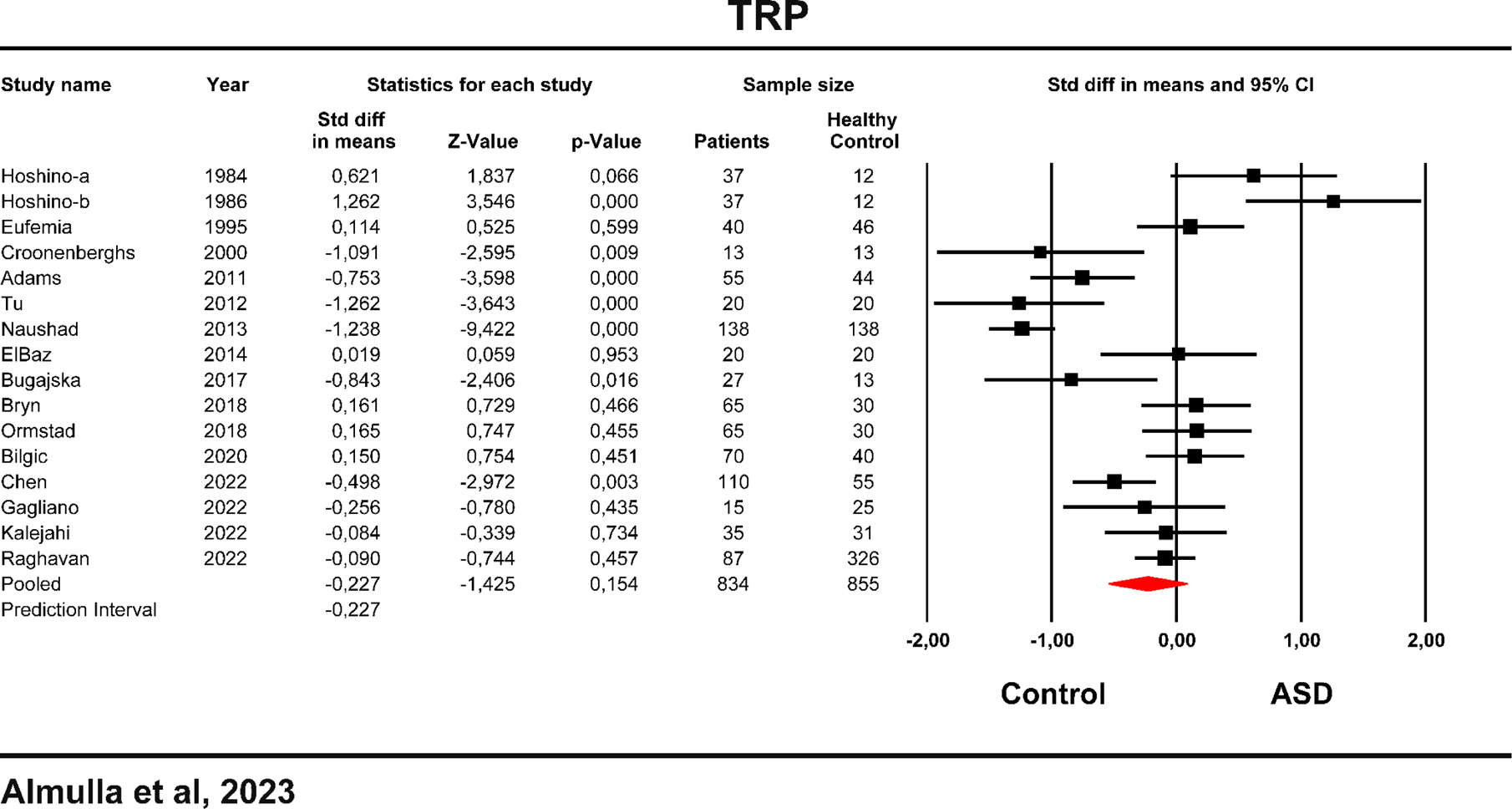
The forest plot of tryptophan (TRP) between patients with autism spectrum disorder (ASD) and healthy controls.

#### KYN/TRP ratio and KYN levels in ASD

The results of the KYN/TRP ratio were obtained from 3 studies. Table 2 indicates no significant alteration in KYN/TRP ratio in ASD (see **Figure 5**), and Table 3 indicates that there was no bias. We examined blood levels of KYN in the present meta-analysis by including 4 studies (see Table 1). Table 2 and ESF, Figure 2 show no significant changes in blood KYN levels in ASD compared to controls. Nevertheless, publication bias analysis displays 1 missing study on the right side of the funnel plot and imputing this study leads to an increased SMD value, indicating a significant elevation of KYN in ASD.

**Figure 5:**
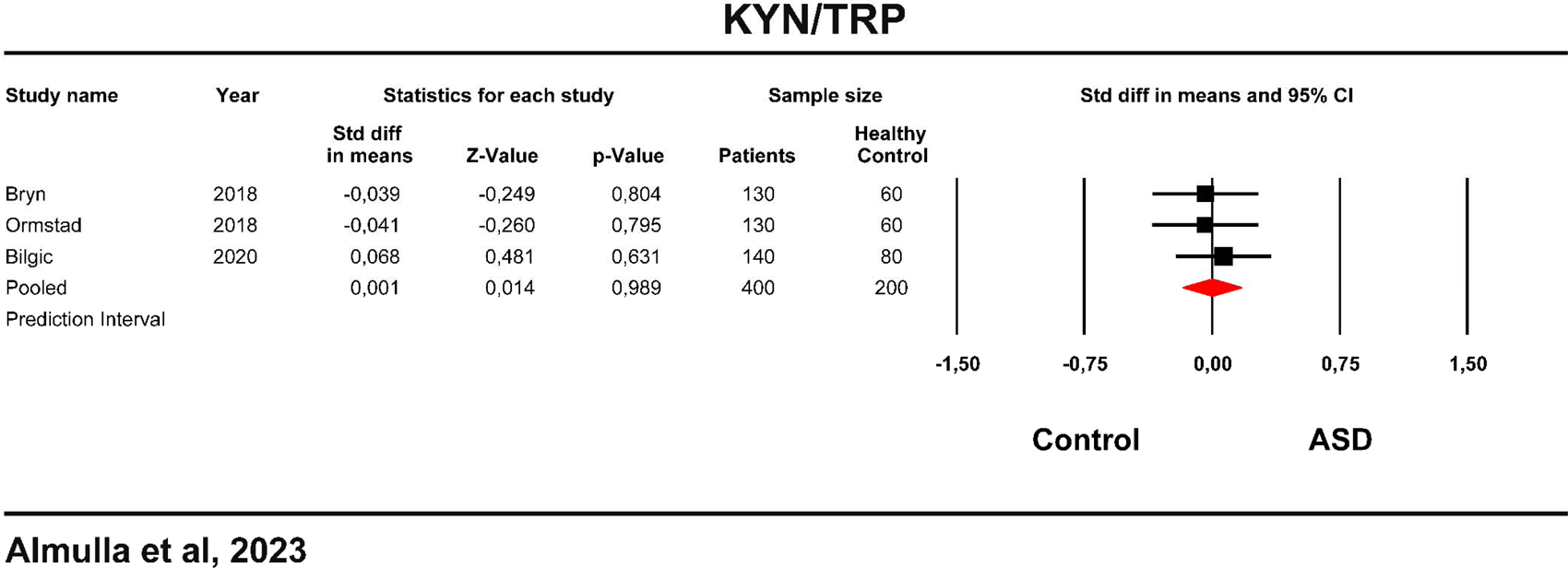
The forest plot of kynurenine (KYN)/tryptophan (TRP) ratio in autism spectrum disorder (ASD) patients and healthy controls.

Urinary KYN levels were also examined in the present study by 2 studies. The results in Table 1 and Table 2 show that patients with ASD have significantly decreased urinary KYN levels compared to healthy controls, with a large effect size (SMD = −1.021).

### Secondary outcome variables

#### KA and the KA/KYN ratio

The results of the KA/KYN ratio were obtained from 5 studies. Table 1 and 2 and ESF, Figure 3, show no significant difference in KA/KYN ratio in ASD compared to healthy controls. Table 3 shows bias with 2 missing studies on the right side of the funnel plot. Adjusting for these studies did not change the results.

We obtained the results of KA levels from 5 studies, as shown in Table 1. Table 2 and ESF, Figure 4 indicate that KA was not altered in ASD. Publication bias analysis revealed 1 missing study on the left side of the funnel plot; adjusting the effect size for this study decreased the SMD value which remained non-significant. Table 1 and 2 indicate no significant change in urinary KA levels.

#### The neurotoxicity index in ASD

The composite score (KYN + 3HK + XA + QA + PA) was obtained from 4 studies, as shown in Table 1 and ESF, Figure 5. The neurotoxicity composite score was not significantly altered in ASD. Table 3 indicates no bias.

### Meta-regression analysis

ESF, Table 6 shows the results of the meta-regression analysis that was conducted to delineate the heterogeneity sources in TRP, CAAs and TRYCATs. Female gender had a substantial negative influence on the results of TRP/CAAs, CAAs, KYN and KA. In addition, sample size also exerted a significant inverse effect on the TRP/CAA ratio and KYN while it positively affected the KA/KYN ratio. Other relevant predictors are shown in ESF, Table 6.

## Discussion

The present study is the first meta-analysis to compare individuals with ASD and healthy controls in terms of a) serum/plasma/urine TRP levels and TRP/CAAs ratio, b) IDO and KAT enzymes activities, and c) solitary TRYCATs including KA.

### Peripheral TRP levels and availability of TRP to the brain

The first major finding of the present study is that patients with ASD show no significant alterations in blood or urine TRP and CAA levels or in TRP/CAA ratio as compared to healthy controls. Similar findings were also reported in preceding studies (D’Eufemia, Finocchiaro et al. 1995, Croonenberghs, Delmeire et al. 2000, ElBaz, Zaki et al. 2014, Bugajska, Berska et al. 2017, Ormstad, Bryn et al. 2018, Kalejahi, Kheirouri et al. 2022). However, some authors found a significantly decreased TRP and TRP/CAA ratio in ASD (Croonenberghs, Delmeire et al. 2000, Naushad, Jain et al. 2013).

Because there are no changes in TRP and in CAA, which compete for transport via the large neutral amino acid transporter 1 (LAT 1) in the BBB (Fernstrom, Larin et al. 1973, Pardridge 1979), the current results suggest no alteration in the availability of TRP to the brain. However, some studies, reported central serotonergic hypoactivity in ASD, as assessed using neuroendocrine studies using 5-hydroxytryptophan, the direct precursor of 5-HT (Croonenberghs, Wauters et al. 2007). While some TRP is present in the blood in a free form, most of this amino acid is weakly bound to serum albumin (Maes, Wauters et al. 1996). The amount of TRP accessible to the brain depends on the levels of albumin, the CAA, and free and total TRP in the serum (Yuwiler, Oldendorf et al. 1977, Pardridge 1979). This is because TRP is transported over the BBB while being stripped of albumin by the capillaries in the BBB (Almulla, Vasupanrajit et al. 2022). Therefore, future research on TRP availability to the brain in ASD should examine free and total TRP levels in association with albumin and CAA levels.

Our findings also revealed no significant increase in the excretion of urine TRP. Previous studies were inconsistent in this regard. For example, some studies found that the excretion of TRP is significantly decreased in ASD patients (Kałuzna-Czaplinska, Michalska et al. 2010, Li, Shen et al. 2018), whereas other studies reported a significantly increased urinary excretion of TRP (Noto, Fanos et al. 2014, Lussu, Noto et al. 2017, Liang, Xiao et al. 2020).

### IDO enzyme and TRYCAY pathway activity in ASD

The second major finding of the current study is that the KYN/TRP ratio in ASD patients was not significantly different from that observed in healthy controls, suggesting normal activity of the IDO enzyme. Some previous studies reported no changes in IDO enzyme activity (Bryn, Verkerk et al. 2017, Ormstad, Bryn et al. 2018, Bilgiç, Abuşoğlu et al. 2022).

Nevertheless, our results show that KYN levels could be increased in ASD after adjusting for possible publication bias. KYN levels in ASD have been reported to be normal in several prior studies (Bryn, Verkerk et al. 2017, Ormstad, Bryn et al. 2018, Bilgiç, Abuşoğlu et al. 2022). On the other hand, our study found significantly decreased excretion of KYN in urine (Gevi, Zolla et al. 2016, Timperio, Gevi et al. 2022). It has been reported that urinary levels of KYN may be decreased in ASD (Gevi, Zolla et al. 2016). Gevi et al found a significant increase in bacterial TRP metabolites, including indolyl 3-acetic acid, indoxyl sulfate and indolyl lactate (Gevi, Zolla et al. 2016). As such, future research should examine plasma/serum as well as urinary KYN and other TRYCATs in the same ASD patients and controls.

Numerous studies have shown that ASD is accompanied by activated peripheral and central immune-inflammatory and nitro-oxidative pathways that may induce IDO enzyme activity (Di Marco, Bonaccorso et al. 2016, Bryn, Aass et al. 2017, Ormstad, Bryn et al. 2018, Bjørklund, Meguid et al. 2020, Pangrazzi, Balasco et al. 2020). As in ASD, previous meta-analyses did not reveal significantly increased IDO enzyme activity in affective disorders (Almulla, Supasitthumrong et al. 2022, Almulla, Thipakorn et al. 2022, Almulla, Thipakorn et al. 2022), although IDO was significantly stimulated in acute COVID-19 infection (Almulla, Supasitthumrong et al. 2022). Probably, the acute inflammatory response in acute COVID-19 is sufficient to stimulate IDO, whereas the chronic mild inflammation in affective disorders and ASD is probably insufficient to trigger IDO (Almulla, Supasitthumrong et al. 2022).

### Neurotoxic TRYCATs in ASD

The third major finding of the current study is that there is no increase in neurotoxic TRYCATs in ASD as assessed with the (KYN + 3HK + XA + QA + PA) composite, the KA/KYN ratio (reflecting KAT enzyme activity) and KA, a neuroprotective TRYCAT. Contrary to the present study, Bryn et found a significantly decreased KA/KYN ratio in ASD (Bryn, Verkerk et al. 2017), although Lim et al. (Lim, Essa et al. 2016) did not find a significant difference in KA between patients and controls. Bilgic et al. showed unaltered KAT enzyme activity although these authors reported significant increased KA levels in ASD (Bilgiç, Abuşoğlu et al. 2022). It should be noted that urinary levels of KA were found to be decreased (Gevi, Zolla et al. 2016) or unaltered (Timperio, Gevi et al. 2022) in ASD. Some studies found a significant elevation in serum and urinary levels of the neurotoxic XA and QA (Gevi, Zolla et al. 2016, Lim, Essa et al. 2016), while other studies revealed no significant difference in QA between ASD and controls (Bryn, Verkerk et al. 2017, Ormstad, Bryn et al. 2018). Another study conducted on platelet-rich plasma found no significant difference in QA between the study groups (Carpita, Nardi et al. 2022).

We recently examined this neurotoxicity index in affective disorders and their severe phenotypes, namely melancholia, psychotic depression, and suicidal behaviors, and these results indicate no significant difference in those patients compared to healthy controls (Almulla, Thipakorn et al. 2022, Almulla, Thipakorn et al. 2022). As discussed above, the chronic mild inflammatory response in those disorders is probably insufficient to cause IDO stimulation, and consequently increased TRYCAT levels.

## Limitations

First, we could not assess the status of the entire TRYCAT pathway along with kynurenine monooxygenase (KMO) enzyme activity, because a limited number of studies are available that evaluated all TRYCATs in ASD. Therefore, additional studies are required to investigate downstream TRYCATs. Second, there could be discrepant results among central and peripheral TRYCATs as detected in schizophrenia, with increased IDO and KAT enzyme activities in the CNS but not in peripheral blood (Almulla, Vasupanrajit et al. 2022). However, the central levels of TRP and TRYCATs could not be assessed in our study since no studies were found on cerebrospinal fluid or brain tissue TRYCATs. We encourage researchers to perform TRP and TRYCAT studies on CSF of ASD patients. Third, the influence of psychotropic drugs could not be examined because only a few studies reported treatment data. Fourth, variations in ASD phenotypes could explain the substantial discrepancies in TRP, TRP/CAAs, and TRYCAT results among ASD patients (Ormstad, Bryn et al. 2018). Hence, it is imperative to diagnose ASD patients in future studies according to the specific ASD phenotypes. Fifth, as discussed above, future research should examine free and total TRP in association with albumin and CAA levels, plasma/serum, and urine KYN, and other TRYCATs in the same ASD patients and controls.

## Conclusions

The present study suggests that there are no aberrations in TRP metabolism and that the availability of TRP to the brain is not affected by increased IDO activity in ASD. Lowered TRP availability and increased neurotoxic TRYCATs do not play a major role in ASD.

## Data Availability

The last author (MM) will reply to reasonable requests for the dataset used in the current meta-analysis after it has been fully utilized by all authors. Excel file will be used to present this dataset.

## Ethical approval and consent to participate

Not applicable.

## Consent for publication

Not applicable.

## Author’s contributions

AA and MM carried out the current study’s design. The data was gathered by YT and AA. The statistical evaluation was done by AA and MM. Each author contributed to the writing and rewriting of the work, and they have all given their consent for submission of the completed version.

## Acknowledgments

Not applicable.

## References

Adams, J. B., T. Audhya, S. McDonough-Means, R. A. Rubin, D. Quig, E. Geis, E. Gehn, M. Loresto, J. Mitchell, S. Atwood, S. Barnhouse and W. Lee (2011). “Nutritional and metabolic status of children with autism vs. neurotypical children, and the association with autism severity.” Nutr Metab (Lond) 8(1): 34.

Almulla, A. F. and M. Maes (2022). “The Tryptophan Catabolite or Kynurenine Pathway’s Role in Major Depression.” Curr Top Med Chem 22(21): 1731–1735.

Almulla, A. F., T. Supasitthumrong, A. Amrapala, C. Tunvirachaisakul, A. K. A. Jaleel, G. Oxenkrug, H. K. Al-Hakeim and M. Maes (2022). “The Tryptophan Catabolite or Kynurenine Pathway in Alzheimer’s Disease: A Systematic Review and Meta-Analysis.” J Alzheimers Dis 88(4): 1325–1339.

Almulla, A. F., T. Supasitthumrong, C. Tunvirachaisakul, A. A. A. Algon, H. K. Al-Hakeim and M. Maes (2022). “The tryptophan catabolite or kynurenine pathway in COVID-19 and critical COVID-19: a systematic review and meta-analysis.” BMC Infectious Diseases 22(1): 615.

Almulla, A. F., Y. Thipakorn, A. A. A. Algon, C. Tunvirachaisakul, H. K. Al-Hakeim and M. Maes (2023). “Reverse cholesterol transport and lipid peroxidation biomarkers in major depression and bipolar disorder: a systematic review and meta-analysis.” medRxiv: 2023.2003.2020.23287483.

Almulla, A. F., Y. Thipakorn, A. Vasupanrajit, A. A. Abo Algon, C. Tunvirachaisakul, A. A. Hashim Aljanabi, G. Oxenkrug, H. K. Al-Hakeim and M. Maes (2022). “The tryptophan catabolite or kynurenine pathway in major depressive and bipolar disorder: A systematic review and meta-analysis.” Brain, Behavior, & Immunity - Health 26: 100537.

Almulla, A. F., Y. Thipakorn, A. Vasupanrajit, C. Tunvirachaisakul, G. Oxenkrug, H. K. Al-Hakeim and M. Maes (2022) “The Tryptophan Catabolite or Kynurenine Pathway in a Major Depressive Episode with Melancholia, Psychotic Features and Suicidal Behaviors: A Systematic Review and Meta-Analysis.” Cells 11 DOI: 10.3390/cells11193112.

Almulla, A. F., A. Vasupanrajit, C. Tunvirachaisakul, H. K. Al-Hakeim, M. Solmi, R. Verkerk and M. Maes (2022). “The tryptophan catabolite or kynurenine pathway in schizophrenia: meta-analysis reveals dissociations between central, serum, and plasma compartments.” Molecular Psychiatry 27(9): 3679–3691.

American Psychiatric Association, D. (2013). Diagnostic and statistical manual of mental disorders: DSM-5, American psychiatric association Washington, DC.

Andrés-Rodríguez, L., X. Borràs, A. Feliu-Soler, A. Pérez-Aranda, N. Angarita-Osorio, P. Moreno-Peral, J. Montero-Marin, J. García-Campayo, A. F. Carvalho, M. Maes and J. V. Luciano (2020). “Peripheral immune aberrations in fibromyalgia: A systematic review, meta-analysis and meta-regression.” Brain, Behavior, and Immunity 87: 881–889.

Ashwood, P. (2023) “Preliminary Findings of Elevated Inflammatory Plasma Cytokines in Children with Autism Who Have Co-Morbid Gastrointestinal Symptoms.” Biomedicines 11 DOI: 10.3390/biomedicines11020436.

Bilgiç, A., S. Abuşoğlu, Ç. Sadıç Çelikkol, M. B. Oflaz, Ö. F. Akça, A. Sivrikaya, T. Baysal and A. Ünlü (2022). “Altered kynurenine pathway metabolite levels in toddlers and preschool children with autism spectrum disorder.” International Journal of Neuroscience 132(8): 826–834.

Bjørklund, G., N. A. Meguid, M. A. El-Bana, A. A. Tinkov, K. Saad, M. Dadar, M. Hemimi, A. V. Skalny, B. Hosnedlová, R. Kizek, J. Osredkar, M. A. Urbina, T. Fabjan, A. A. El-Houfey, J. Kałużna-Czaplińska, P. Gątarek and S. Chirumbolo (2020). “Oxidative Stress in Autism Spectrum Disorder.” Mol Neurobiol 57(5): 2314–2332.

Bryn, V., H. C. Aass, O. H. Skjeldal, J. Isaksen, O. D. Saugstad and H. Ormstad (2017). “Cytokine Profile in Autism Spectrum Disorders in Children.” J Mol Neurosci 61(1): 1–7.

Bryn, V., R. Verkerk, O. H. Skjeldal, O. D. Saugstad and H. Ormstad (2017). “Kynurenine Pathway in Autism Spectrum Disorders in Children.” Neuropsychobiology 76(2): 82–88.

Bugajska, J., J. Berska, T. Wojtyto, M. Bik-Multanowski and K. Sztefko (2017). “The amino acid profile in blood plasma of young boys with autism.” Psychiatria Polska 51(2): 359–368.

Carpita, B., B. Nardi, L. Palego, I. M. Cremone, G. Massimetti, C. Carmassi, L. Betti, G. Giannaccini and L. Dell’Osso (2022). “Kynurenine pathway and autism spectrum phenotypes: an investigation among adults with autism spectrum disorder and their first-degree relatives.” CNS Spectr: 1–12.

Chauhan, A., V. Chauhan, W. T. Brown and I. Cohen (2004). “Oxidative stress in autism: increased lipid peroxidation and reduced serum levels of ceruloplasmin and transferrin--the antioxidant proteins.” Life Sci 75(21): 2539–2549.

Chauhan, A., F. Gu, M. M. Essa, J. Wegiel, K. Kaur, W. T. Brown and V. Chauhan (2011). “Brain region-specific deficit in mitochondrial electron transport chain complexes in children with autism.” J Neurochem 117(2): 209–220.

Chen, W. X., Y. R. Chen, M. Z. Peng, X. Liu, Y. N. Cai, Z. F. Huang, S. Y. Yang, J. Y. Huang, R. H. Wang, P. Yi and L. Liu (2023). “Plasma Amino Acid Profile in Children with Autism Spectrum Disorder in Southern China: Analysis of 110 Cases.” J Autism Dev Disord.

Cohen, J. (2013). Statistical power analysis for the behavioral sciences, Academic press.

Corbett, B. A., C. W. Schupp, D. Simon, N. Ryan and S. Mendoza (2010). “Elevated cortisol during play is associated with age and social engagement in children with autism.” Molecular Autism 1(1): 13.

Croonenberghs, J., E. Bosmans, D. Deboutte, G. Kenis and M. Maes (2002). “Activation of the inflammatory response system in autism.” Neuropsychobiology 45(1): 1–6.

Croonenberghs, J., L. Delmeire, R. Verkerk, A. H. Lin, A. Meskal, H. Neels, M. Van der Planken, S. Scharpe, D. Deboutte, G. Pison and M. Maes (2000). “Peripheral markers of serotonergic and noradrenergic function in post-pubertal, caucasian males with autistic disorder.” Neuropsychopharmacology 22(3): 275–283.

Croonenberghs, J., R. Verkerk, S. Scharpe, D. Deboutte and M. Maes (2005). “Serotonergic disturbances in autistic disorder: L-5-hydroxytryptophan administration to autistic youngsters increases the blood concentrations of serotonin in patients but not in controls.” Life Sci 76(19): 2171–2183.

Croonenberghs, J., A. Wauters, D. Deboutte, R. Verkerk, S. Scharpe and M. Maes (2007). “Central serotonergic hypofunction in autism: results of the 5-hydroxy-tryptophan challenge test.” Neuroendocrinology Letters 28(4): 449–455.

D’Eufemia, P., R. Finocchiaro, M. Celli, L. Viozzi, D. Monteleone and O. Giardini (1995). “Low serum tryptophan to large neutral amino acids ratio in idiopathic infantile autism.” Biomed Pharmacother 49(6): 288–292.

Di Marco, B., C. M. Bonaccorso, E. Aloisi, S. D’Antoni and M. V. Catania (2016). “Neuro-Inflammatory Mechanisms in Developmental Disorders Associated with Intellectual Disability and Autism Spectrum Disorder: A Neuro-Immune Perspective.” CNS Neurol Disord Drug Targets 15(4): 448–463.

Duval, S. and R. Tweedie (2000). “A nonparametric “trim and fill” method of accounting for publication bias in meta-analysis.” Journal of the american statistical association 95(449): 89–98.

ElBaz, F. M., M. M. Zaki, A. M. Youssef, G. F. ElDorry and D. Y. Elalfy (2014). “Study of plasma amino acid levels in children with autism: An Egyptian sample.” Egyptian Journal of Medical Human Genetics 15(2): 181–186.

Fernstrom, J. D., F. Larin and R. J. Wurtman (1973). “Correlation between brain tryptophan and plasma neutral amino acid levels following food consumption in rats.” Life Sciences 13(5): 517–524.

Fombonne, E. (2018). “Editorial: The rising prevalence of autism.” Journal of Child Psychology and Psychiatry 59(7): 717–720.

Gabriele, S., R. Sacco and A. M. Persico (2014). “Blood serotonin levels in autism spectrum disorder: a systematic review and meta-analysis.” Eur Neuropsychopharmacol 24(6): 919–929.

Gagliano, A., F. Murgia, A. M. Capodiferro, M. G. Tanca, A. Hendren, S. G. Falqui, M. Aresti, M. Comini, S. Carucci, E. Cocco, L. Lorefice, M. Roccella, L. Vetri, S. Sotgiu, A. Zuddas and L. Atzori (2022). “(1)H-NMR-Based Metabolomics in Autism Spectrum Disorder and Pediatric Acute-Onset Neuropsychiatric Syndrome.” J Clin Med 11(21).

Gardner, R. M., B. K. Lee, M. Brynge, H. Sjöqvist, C. Dalman and H. Karlsson (2021). “Neonatal Levels of Acute Phase Proteins and Risk of Autism Spectrum Disorder.” Biological Psychiatry 89(5): 463–475.

Gevi, F., L. Zolla, S. Gabriele and A. M. Persico (2016). “Urinary metabolomics of young Italian autistic children supports abnormal tryptophan and purine metabolism.” Mol Autism 7: 47.

Hergüner, S., F. M. Keleşoğlu, C. Tanıdır and M. Çöpür (2012). “Ferritin and iron levels in children with autistic disorder.” European Journal of Pediatrics 171(1): 143–146.

Higgins, J. P., J. Thomas, J. Chandler, M. Cumpston, T. Li, M. J. Page and V. A. Welch (2019). Cochrane handbook for systematic reviews of interventions, John Wiley & Sons.

Hoshino, Y., T. Yamamoto, M. Kaneko and H. Kumashiro (1986). “Plasma free tryptophan concentration in autistic children.” Brain Dev 8(4): 424–427.

Hoshino, Y., T. Yamamoto, M. Kaneko, R. Tachibana, M. Watanabe, Y. Ono and H. Kumashiro (1984). “Blood serotonin and free tryptophan concentration in autistic children.” Neuropsychobiology 11(1): 22–27.

Kalejahi, P., S. Kheirouri and S. G. Noorazar (2022). “Comparison of blood tryptophan and glutamate levels in children and adolescents with autism and healthy controls.” Cognition, Brain, Behavior 26(2): 89–99.

Kałuzna-Czaplinska, J., M. Michalska and J. Rynkowski (2010). “Determination of tryptophan in urine of autistic and healthy children by gas chromatography/mass spectrometry.” Med Sci Monit 16(10): Cr488–492.

Li, C., K. Shen, L. Chu, P. Liu, Y. Song and X. Kang (2018). “Decreased levels of urinary free amino acids in children with autism spectrum disorder.” J Clin Neurosci 54: 45–49.

Liang, Y., Z. Xiao, X. Ke, P. Yao, Y. Chen, L. Lin and J. Lu (2020). “Urinary Metabonomic Profiling Discriminates Between Children with Autism and Their Healthy Siblings.” Med Sci Monit 26: e926634.

Lim, C. K., M. M. Essa, R. de Paula Martins, D. B. Lovejoy, A. A. Bilgin, M. I. Waly, Y. M. Al-Farsi, M. Al-Sharbati, M. A. Al-Shaffae and G. J. Guillemin (2016). “Altered kynurenine pathway metabolism in autism: Implication for immune-induced glutamatergic activity.” Autism Res 9(6): 621–631.

Liu, X., J. Lin, H. Zhang, N. U. Khan, J. Zhang, X. Tang, X. Cao and L. Shen (2022). “Oxidative Stress in Autism Spectrum Disorder—Current Progress of Mechanisms and Biomarkers.” Frontiers in Psychiatry 13.

Lussu, M., A. Noto, A. Masili, A. C. Rinaldi, A. Dessì, M. De Angelis, A. De Giacomo, V. Fanos, L. Atzori and R. Francavilla (2017). “The urinary (1) H-NMR metabolomics profile of an italian autistic children population and their unaffected siblings.” Autism Res 10(6): 1058–1066.

Maes, M., B. E. Leonard, A. M. Myint, M. Kubera and R. Verkerk (2011). “The new ‘5-HT’ hypothesis of depression: Cell-mediated immune activation induces indoleamine 2,3-dioxygenase, which leads to lower plasma tryptophan and an increased synthesis of detrimental tryptophan catabolites (TRYCATs), both of which contribute to the onset of depression.” Progress in Neuro-Psychopharmacology and Biological Psychiatry 35(3): 702–721.

Maes, M., A. Wauters, R. Verkerk, P. Demedts, H. Neels, A. Van Gastel, P. Cosyns, S. Scharpé and R. Desnyder (1996). “Lower serum L-tryptophan availability in depression as a marker of a more generalized disorder in protein metabolism.” Neuropsychopharmacology 15(3): 243–251.

Manivasagam, T., S. Arunadevi, M. M. Essa, C. SaravanaBabu, A. Borah, A. J. Thenmozhi and M. W. Qoronfleh (2020). “Role of oxidative stress and antioxidants in autism.” Personalized Food Intervention and Therapy for Autism Spectrum Disorder Management: 193–206.

Masi, A., D. S. Quintana, N. Glozier, A. R. Lloyd, I. B. Hickie and A. J. Guastella (2015). “Cytokine aberrations in autism spectrum disorder: a systematic review and meta-analysis.” Molecular Psychiatry 20(4): 440–446.

Nadeem, A., S. F. Ahmad, L. Y. Al-Ayadhi, S. M. Attia, N. O. Al-Harbi, K. S. Alzahrani and S. A. Bakheet (2020). “Differential regulation of Nrf2 is linked to elevated inflammation and nitrative stress in monocytes of children with autism.” Psychoneuroendocrinology 113: 104554.

Nadeem, R., T. Hussain and H. Sajid (2020). “C reactive protein elevation among children or among mothers’ of children with autism during pregnancy, a review and meta-analysis.” BMC Psychiatry 20(1): 251.

Naushad, S. M., J. M. Jain, C. K. Prasad, U. Naik and R. R. Akella (2013). “Autistic children exhibit distinct plasma amino acid profile.” Indian J Biochem Biophys 50(5): 474–478.

Nie, Z.-Q., D. Han, K. Zhang, M. Li, H.-K. Kwon, S.-H. Im, L. Xu, J.-c. Yang, Z.-W. Li, X.-W. Huang, J. Wen, Y. Shu-Jun, F. Yin, C. Shen, P. Ashwood, C.-Y. Kang and X. Cao (2023). “TH1/Treg ratio may be a marker of autism in children with immune dysfunction.” Research in Autism Spectrum Disorders 101: 102085.

Noto, A., V. Fanos, L. Barberini, D. Grapov, C. Fattuoni, M. Zaffanello, A. Casanova, G. Fenu, A. De Giacomo, M. De Angelis, C. Moretti, P. Papoff, R. Ditonno and R. Francavilla (2014). “The urinary metabolomics profile of an Italian autistic children population and their unaffected siblings.” J Matern Fetal Neonatal Med 27 Suppl 2: 46–52.

Olesova, D., J. Galba, J. Piestansky, H. Celusakova, G. Repiska, K. Babinska, D. Ostatnikova, S. Katina and A. Kovac (2020). “A Novel UHPLC-MS Method Targeting Urinary Metabolomic Markers for Autism Spectrum Disorder.” Metabolites 10(11).

Ormstad, H., V. Bryn, O. D. Saugstad, O. Skjeldal and M. Maes (2018). “Role of the Immune System in Autism Spectrum Disorders (ASD).” CNS & Neurological Disorders - Drug Targets-CNS & Neurological Disorders) 17(7): 489–495.

Ormstad, H., V. Bryn, R. Verkerk, O. H. Skjeldal, B. Halvorsen, O. D. Saugstad, J. Isaksen and M. Maes (2018). “Serum Tryptophan, Tryptophan Catabolites and Brain-derived Neurotrophic Factor in Subgroups of Youngsters with Autism Spectrum Disorders.” CNS Neurol Disord Drug Targets 17(8): 626–639.

Ou, J. J., L. J. Shi, G. L. Xun, C. Chen, R. R. Wu, X. R. Luo, F. Y. Zhang and J. P. Zhao (2015). “Employment and financial burden of families with preschool children diagnosed with autism spectrum disorders in urban China: results from a descriptive study.” BMC Psychiatry 15: 3.

Page, M. J., J. E. McKenzie, P. M. Bossuyt, I. Boutron, T. C. Hoffmann, C. D. Mulrow, L. Shamseer, J. M. Tetzlaff, E. A. Akl and S. E. Brennan (2021). “The PRISMA 2020 statement: an updated guideline for reporting systematic reviews.” International journal of surgery 88: 105906.

Pangrazzi, L., L. Balasco and Y. Bozzi (2020). “Oxidative Stress and Immune System Dysfunction in Autism Spectrum Disorders.” Int J Mol Sci 21(9).

Pardridge, W. M. (1979). “Tryptophan transport through the blood-brain barrier: In vivo measurement of free and albumin-bound amino acid.” Life Sciences 25(17): 1519–1528.

Picardi, A., A. Gigantesco, E. Tarolla, V. Stoppioni, R. Cerbo, M. Cremonte, G. Alessandri, I. Lega and F. Nardocci (2018). “Parental Burden and its Correlates in Families of Children with Autism Spectrum Disorder: A Multicentre Study with Two Comparison Groups.” Clin Pract Epidemiol Ment Health 14: 143–176.

Raghavan, R., N. S. Anand, G. Wang, X. Hong, C. Pearson, B. Zuckerman, H. Xie and X. Wang (2022). “Association between cord blood metabolites in tryptophan pathway and childhood risk of autism spectrum disorder and attention-deficit hyperactivity disorder.” Transl Psychiatry 12(1): 270.

Saghazadeh, A., B. Ataeinia, K. Keynejad, A. Abdolalizadeh, A. Hirbod-Mobarakeh and N. Rezaei (2019). “Anti-inflammatory cytokines in autism spectrum disorders: A systematic review and meta-analysis.” Cytokine 123: 154740.

Spratt, E. G., J. S. Nicholas, K. T. Brady, L. A. Carpenter, C. R. Hatcher, K. A. Meekins, R. W. Furlanetto and J. M. Charles (2012). “Enhanced cortisol response to stress in children in autism.” J Autism Dev Disord 42(1): 75–81.

Timperio, A. M., F. Gevi, F. Cucinotta, A. Ricciardello, L. Turriziani, M. L. Scattoni and A. M. Persico (2022). “Urinary Untargeted Metabolic Profile Differentiates Children with Autism from Their Unaffected Siblings.” Metabolites 12(9).

Tripathi, M. K., M. Kartawy and H. Amal (2020). “The role of nitric oxide in brain disorders: Autism spectrum disorder and other psychiatric, neurological, and neurodegenerative disorders.” Redox Biology 34: 101567.

Tu, W. J., H. Chen and J. He (2012). “Application of LC-MS/MS analysis of plasma amino acids profiles in children with autism.” J Clin Biochem Nutr 51(3): 248–249.

Wan, X., W. Wang, J. Liu and T. Tong (2014). “Estimating the sample mean and standard deviation from the sample size, median, range and/or interquartile range.” BMC medical research methodology 14: 1–13.

Wang, J., B. Ma, J. Wang, Z. Zhang and O. Chen (2022). “Global prevalence of autism spectrum disorder and its gastrointestinal symptoms: A systematic review and meta-analysis.” Frontiers in Psychiatry 13.

Warren, R. P., N. C. Margaretten, N. C. Pace and A. Foster (1986). “Immune abnormalities in patients with autism.” Journal of Autism and Developmental Disorders 16(2): 189–197.

Wing, L. and J. Gould (1979). “Severe impairments of social interaction and associated abnormalities in children: epidemiology and classification.” J Autism Dev Disord 9(1): 11–29.

Xu, N., X. Li and Y. Zhong (2015). “Inflammatory Cytokines: Potential Biomarkers of Immunologic Dysfunction in Autism Spectrum Disorders.” Mediators of Inflammation 2015: 531518.

Yenkoyan, K., A. Grigoryan, K. Fereshetyan and D. Yepremyan (2017). “Advances in understanding the pathophysiology of autism spectrum disorders.” Behavioural Brain Research 331: 92–101.

Yin, F., H. Wang, Z. Liu and J. Gao (2020). “Association between peripheral blood levels of C-reactive protein and Autism Spectrum Disorder in children: A systematic review and meta-analysis.” Brain Behav Immun 88: 432–441.

Yuwiler, A., W. H. Oldendorf, E. Geller and L. Braun (1977). “Effect of albumin binding and amino acid competition on tryptophan uptake into brain.” J Neurochem 28(5): 1015–1023.

Zhao, H., H. Zhang, S. Liu, W. Luo, Y. Jiang and J. Gao (2021). “Association of Peripheral Blood Levels of Cytokines With Autism Spectrum Disorder: A Meta-Analysis.” Frontiers in Psychiatry 12.

