## supplementary file for "The tryptophan catabolite or kynurenine pathway in autism spectrum disorder; a systematic review and meta-analysis"

SHORT TITLE: Kynurenine pathway in autism spectrum disorder

Abbas F. Almulla, Ph.D.^a,b^ , Yanin Thipakorn, M.D., Ph.D.^a^ , Chavit Tunvirachaisakul, M.D., Ph.D.^a,c^ , Michael Maes, M.D., Ph.D.^a,c,d,e,f^

^a^ Department of Psychiatry, Faculty of Medicine, Chulalongkorn University, Bangkok, Thailand.

^b^ Medical Laboratory Technology Department, College of Medical Technology, The Islamic University, Najaf, Iraq.

^c^ Cognitive Impairment and Dementia Research Unit, Faculty of Medicine, Chulalongkorn University, Bangkok, Thailand.

^d^ Department of Psychiatry, Medical University of Plovdiv, Plovdiv, Bulgaria.

^e^. Research Institute, Medical University of Plovdiv, Plovdiv, Bulgaria.

^f^ Kyung Hee University, 26 Kyungheedae-ro, Dongdaemun-gu, Seoul 02447, Korea

**Corresponding author:**

Prof. Dr Michael Maes, M.D., Ph.D.

Department of Psychiatry

Faculty of Medicine, Chulalongkorn University

Bangkok, 10330

Thailand

E-mail addresses:

**ESF, Table 1.** Search sentences and terms were used in each database

| **Database Name** | **Search Sentence** | **No. of Articles** |
| --- | --- | --- |
| **PubMed/Medline** | (("Autistic Disorder"[MeSH] OR autism[Title/Abstract]) AND ("Tryptophan"[MeSH] OR tryptophan[Title/Abstract] OR "Kynurenine"[MeSH] OR kynurenine[Title/Abstract] OR "Kynurenic Acid"[MeSH] OR kynurenic acid[Title/Abstract] OR "Quinolinic Acid"[MeSH] OR quinolinic acid[Title/Abstract] OR "Xanthurenic Acid"[MeSH] OR xanthurenic acid[Title/Abstract] OR "3-Hydroxykynurenine"[MeSH] OR hydroxykynurenine[Title/Abstract] OR "Anthranilic Acids"[MeSH] OR anthranilic acid[Title/Abstract] OR "Picolinic Acids"[MeSH] OR picolinic acid[Title/Abstract])) | **244** |
|  | (("ASD"[MeSH] OR autism[Title/Abstract]) AND ("TRP"[MeSH] OR TRY[Title/Abstract] OR "KYN"[MeSH] OR KYN[Title/Abstract] OR "KA"[MeSH] OR KYNA[Title/Abstract] OR "QA"[MeSH] OR QUINA[Title/Abstract] OR "XA"[MeSH] OR XAN[Title/Abstract] OR "3-HK"[MeSH] OR OHKYNTitle/Abstract] OR "AA"[MeSH] OR ANA[Title/Abstract] OR "PA"[MeSH] OR pico[Title/Abstract])) | **1581** |
| **Google Scholar** | autism AND (tryptophan OR kynurenine OR "kynurenic acid" OR "quinolinic acid" OR "xanthurenic acid" OR hydroxykynurenine OR "anthranilic acid" OR "picolinic acid") | **26000** |
|  | Autism spectrum disorders AND (tryptophan OR kynurenine OR "kynurenic acid" OR "quinolinic acid" OR "xanthurenic acid" OR hydroxykynurenine OR "anthranilic acid" OR "picolinic acid") | **18700** |
| **SciFinder** | autism AND (tryptophan OR kynurenine OR kynurenic acid OR quinolinic acid OR xanthurenic acid OR hydroxykynurenine OR anthranilic acid OR picolinic acid) | **2441** |
|  | Autism spectrum disorders AND (tryptophan OR kynurenine OR kynurenic acid OR quinolinic acid OR xanthurenic acid OR hydroxykynurenine OR anthranilic acid OR picolinic acid) | **1426** |

**ESF, Table 2.** Immune cofounder’s scale (ICS) applied from Andrés-Rodríguez, et al., 2019

| **Methodological quality of the study** | |
| --- | --- |
| **1** | Study sample ≥ 128 participants including patients and controls (1= Yes, 0 = No) |
| **2** | Did the study control the results for potential confounders (e.g., age, BMI, gender, race)? (1= Yes, 0 = No) |
| **3** | Were participants with autism spectrum disorder and controls age- and-gender-matched or was there a statistical control? (1= Yes, 0 = No) |
| **4** | Was the time of sample collection specified (e.g., morning vs. evening)? (1= Yes, 0 = No) |
| **5** | Were participants with autism spectrum disorder free of immunomodulatory drugs including anti-cytokines, glucocorticoids, immunoglobulins, and immunosuppressants, or was there a medication washout period, or was drug intake statistically controlled for? (1= Yes, 0 = No) |
| **6** | Were participants with autism spectrum disorder free of antidepressants and mood stabilizers or were the data statistically controlled for? (1= Yes, 0 = No) |
| **7** | Reporting either the manufacturer of the test or detection limit and coefficients of variation (1= Yes, 0 = No) |
| **8** | Reporting how data under detection limit were handled (1 = Yes, 0 = No) |
| **9** | Reporting % of the sample under detection limit (1=Yes, 0= No) |
| **10** | Reporting blood fraction (serum, plasma, culture supernatant or whole blood) (1= Yes, 0 = No) |
| **Total quality score (10 points)** | |
| **Biomarker confounders red points**  *The red points should not be given if the item is statistically controlled for* | |
| **1** | 3 red points for comorbid illnesses such as autoimmune disorders & other immune disorders including rheumatoid arthritis, psoriasis, inflammatory bowel disease, chronic obstructive pulmonary disease, multiple sclerosis |
| **2** | 3 red points for use of recreational drugs such as methamphetamine or opioids |
| **3** | 2 red points when groups were not matched for age |
| **4** | 2 red points when groups were not matched for sex |
| **5** | 2 red points for medication use as for example immunomodulators |
| **6** | 2 red points for early traumatic life events |
| **7** | 2 red points for shift work and primary sleep disorders |
| **8** | 1.5 red points for use of antipsychotics |
| **9** | 1 red point for more common systemic immune disorders including diabetes type 1/2, essential hypertension, metabolic syndrome |
| **10** | 1 red point for not fasting (8 hours before blood extraction) |
| **11** | 1 red point for use of omega-3 and antioxidant supplements |
| **12** | 1 red point when data were not controlled for body mass index |
| **13** | 1 red point when data were not controlled for physical activity or sedentary life |
| **14** | 1 red point when data were not controlled for smoking |
| **15** | 1 red point for use of oral contraceptives or NSAIDs |
| **16** | 0.5 red points when data were not controlled for ethnicity in countries such as US, Brazil |
| **17** | 0.5 red points when data were not controlled for seasonality |
| **18** | 0.5 red points when data were not controlled for diurnal variation (8-10 a.m. versus all other time points) |
|  | **Total red point score (26 points)** |

**ESF, Table 3.** PRISMA checklist

| **Section/topic** | **#** | **Checklist item** | **Reported on page #** |
| --- | --- | --- | --- |
| **TITLE** | | | |
| Title | 1 | Identify the report as a systematic review, meta-analysis, or both. | 1 |
| **ABSTRACT** | | | |
| Structured summary | 2 | Provide a structured summary including, as applicable: background; objectives; data sources; study eligibility criteria, participants, and interventions; study appraisal and synthesis methods; results; limitations; conclusions and implications of key findings; systematic review registration number. | 3,4 |
| **INTRODUCTION** | | | |
| Rationale | 3 | Describe the rationale for the review in the context of what is already known. | 5-7 |
| Objectives | 4 | Provide an explicit statement of questions being addressed with reference to participants, interventions, comparisons, outcomes, and study design (PICOS). | 8 |
| **METHODS** | | | |
| Protocol and registration | 5 | Indicate if a review protocol exists, if and where it can be accessed (e.g., Web address), and, if available, provide registration information including registration number. | 8,9 |
| Eligibility criteria | 6 | Specify study characteristics (e.g., PICOS, length of follow-up) and report characteristics (e.g., years considered, language, publication status) used as criteria for eligibility, giving rationale. | 9 |
| Information sources | 7 | Describe all information sources (e.g., databases with dates of coverage, contact with study authors to identify additional studies) in the search and date last searched. | ESF, Table 1 |
| Search | 8 | Present full electronic search strategy for at least one database, including any limits used, such that it could be repeated. | ESF, Table 1 |
| Study selection | 9 | State the process for selecting studies (i.e., screening, eligibility, included in systematic review, and, if applicable, included in the meta-analysis). | 10,11 |
| Data collection process | 10 | Describe method of data extraction from reports (e.g., piloted forms, independently, in duplicate) and any processes for obtaining and confirming data from investigators. | 11 |
| Data items | 11 | List and define all variables for which data were sought (e.g., PICOS, funding sources) and any assumptions and simplifications made. | 11 |
| Risk of bias in individual studies | 12 | Describe methods used for assessing risk of bias of individual studies (including specification of whether this was done at the study or outcome level), and how this information is to be used in any data synthesis. | 12 |
| Summary measures | 13 | State the principal summary measures (e.g., risk ratio, difference in means). | 12 |
| Synthesis of results | 14 | Describe the methods of handling data and combining results of studies, if done, including measures of consistency (e.g., I^2^) for each meta-analysis. | 12 |
| Risk of bias across studies | 15 | Specify any assessment of risk of bias that may affect the cumulative evidence (e.g., publication bias, selective reporting within studies). | 12 |
| Additional analyses | 16 | Describe methods of additional analyses (e.g., sensitivity or subgroup analyses, meta-regression), if done, indicating which were pre-specified. | 12 |
| **RESULTS** | | |  |
| Study selection | 17 | Give numbers of studies screened, assessed for eligibility, and included in the review, with reasons for exclusions at each stage, ideally with a flow diagram. | 13 and ESF, Table 1 and Table 2 |
| Study characteristics | 18 | For each study, present characteristics for which data were extracted (e.g., study size, PICOS, follow-up period) and provide the citations. | ESF, Table 5 |
| Risk of bias within studies | 19 | Present data on risk of bias of each study and, if available, any outcome level assessment (see item 12). | Table 3 |
| Results of individual studies | 20 | For all outcomes considered (benefits or harms), present, for each study: (a) simple summary data for each intervention group (b) effect estimates and confidence intervals, ideally with a forest plot. | 14,15 |
| Synthesis of results | 21 | Present results of each meta-analysis done, including confidence intervals and measures of consistency. | Table 1 and Table 2 |
| Risk of bias across studies | 22 | Present results of any assessment of risk of bias across studies (see Item 15). | Table 3 |
| Additional analysis | 23 | Give results of additional analyses, if done (e.g., sensitivity or subgroup analyses, meta-regression [see Item 16]). | 16, ESF, Table 6 |
| **DISCUSSION** | | |  |
| Summary of evidence | 24 | Summarize the main findings including the strength of evidence for each main outcome; consider their relevance to key groups (e.g., healthcare providers, users, and policy makers). | 16-20 |
| Limitations | 25 | Discuss limitations at study and outcome level (e.g., risk of bias), and at review-level (e.g., incomplete retrieval of identified research, reporting bias). | 20 |
| Conclusions | 26 | Provide a general interpretation of the results in the context of other evidence, and implications for future research. | 21 |
| **FUNDING** | | |  |
| Funding | 27 | Describe sources of funding for the systematic review and other support (e.g., supply of data); role of funders for the systematic review. | 21 |

**ESF, Table 4.** Studies excluded from the meta-analysis but included in the systematic review.

| **Authors, year** | **Reason why excluded from the meta-analysis** |
| --- | --- |
| (Boccuto, Chen et al. 2013) | No mean(SD) or any other way to extract them |
| (Kałużna-Czaplińska 2011) | Outlier |
| (Minderaa, Anderson et al. 1989) | Whole blood |
| (Minderaa, Anderson et al. 1987) | Whole blood |

**ESF, table 5.** Characteristics of the studies included in the systematic reviews and meta-analysis

| **NO** | **Authors, years** | **Setting** | **Type of case** | **Type of Control** | **Sample Size** | | | **Age** | | **Assessed**  **Biomarkers** | **Specimen** | **Method** | **Quality score** | **Red point score** | **Findnigs** |
| --- | --- | --- | --- | --- | --- | --- | --- | --- | --- | --- | --- | --- | --- | --- | --- |
|  |  |  |  |  | **Cases**  **M/F** | **Control**  **M/F** | **Total**  **M/F** | **Case-Mean (SD)** | **Control- Mean(SD)** |  |  |  |  |  |  |
| 1 | (Hoshino, Yamamoto et al. 1984) | Japan | Autism | Healthy Control | 37(34/3) | 12(6/6) | 49(40/9) | 4.7(0) | 10.5(0) | TRP | Plasma | Chemical | 6,5 | 5,5 | TRP# |
| 2 | (Hoshino, Yamamoto et al. 1986) | Japan | Autism | Healthy Control | 37(34/3) | 12(6/6) | 49(40/9) | 4.7(0) | 10.5(0) | TRP | Plasma | Chemical | 6,5 | 5,5 | TRP# |
| 3 | (Minderaa, Anderson et al. 1987) | USA | Autism | Healthy Control | 16(11/5), 20(16/4) | 27(19/8) | 43(30/13) | 20.6(4.6), 19.4(4.1) | 20.3(6.9) | TRP | Whole blood | HPLC | 5,5 | 5 | TRP* |
| 4 | (Minderaa, Anderson et al. 1989) | USA | Autism | Healthy Control | 40(28/12) | 20(15/5) | 60(43/17) | 19.4(4.9) | 22.0(7.5) | TRP | Whole blood | HPLC | 5,5 | 5 | TRP* |
| 5 | (D'Eufemia, Finocchiaro et al. 1995) | Italy | Autism | Healthy Control | 40(27/13) | 46(27/19) | 86(54/32) | 12.4(0) | 11.2(0) | Isoleucine, Leucine, Phenylalanine,TRP,Tyrosine,Valine | Serum | HPLC | 7,5 | 6 | Isoleucine#, Leucine#, Phenylalanine#,TRP#,Tyrosine#,Valine# |
| 6 | (Croonenberghs, Delmeire et al. 2000) | USA | Autism | Healthy Control | 13(13/0) | 13(13/0) | 26(26/0) | 14.5(1.8) | 15.1(1.5) | CAA, Isoleucine, Leucine, Phenylalanine,TRP,Tyrosine,Valine | Serum | HPLC | 7,5 | 6 | CAA*, Isoleucine*, Leucine*, Phenylalanine*,TRP*,Tyrosine*,Valine* |
| 7 | (Kałuzna-Czaplinska, Michalska et al. 2010) | Poland | Autism | Healthy Control | 10(9/1) | 21(13/8) | 31(22/9) | 0 | 0 | TRP | Urine | GC/MS | 8 | 7,5 | TRP* |
| 8 | (Adams, Audhya et al. 2011) | USA | Autism | neurotypical | 55(49/6) | 44(39/5) | 99(88/11) | 10.0(3.1) | 11.0(3.1) | Isoleucine,Leucine,Phenylalanine,TRP,Tyrosine,Valine | Plasma | HPLC | 5,5 | 6 | Isoleucine*,Leucine(NS),Phenylalanine*,TRP*,Tyrosine*,Valine* |
| 9 | (Kałużna-Czaplińska 2011) | Poland | Autism | Healthy Control | 35(30/5) | 36(28/8) | 71(58/13) | 0 | 0 | TRP | Urine | GC/MS | 5,5 | 6,5 | TRP* |
| 10 | (Tu, Chen et al. 2012) | China | Autism | nonconsecutive control | 20(17/3) | 20(17/3) | 40(34/6) | 3.46(0.56) | 0 | Leucine,TRP,Tyrosine,Valine | Plasma | LC-MS/MS | 5,5 | 6 | Leucine*,TRP*,Tyrosine*,Valine* |
| 11 | (Boccuto, Chen et al. 2013) | USA | Autism | Healthy Control | 87(74/13) | 78(66/12) | 165(140/25) | 2.5(0) | 4.17(0) | TRP | Lymphocytes | MicroArray | 7 | 4,5 | TRP* |
| 12 | (Naushad, Jain et al. 2013) | India | Autism | Healthy Control | 138(120/18) | 138(120/18) | 276(240/36) | 4.4(1.7) | 4.4(1.6) | Isoleucine,Leucine,Phenylalanine,TRP,Tyrosine,Valine | Plasma | HPLC | 8,5 | 6 | Isoleucine*,Leucine#,Phenylalanine*,TRP*,Tyrosine*,Valine# |
| 13 | (ElBaz, Zaki et al. 2014) | Egypt | Autism | Healthy Control | 20(19/1) | 20(9/11) | 40(28/12) | 4.65(1.67) | 4.65(1.67) | Isoleucine,Leucine,Phenylalanine,TRP,Tyrosine,Valine | Plasma | HPLC | 6,5 | 7 | Isoleucine*,Leucine*,Phenylalanine*,TRP#,Tyrosine*,Valine* |
| 14 | (Noto, Fanos et al. 2014) | Italy | Autism | Healthy Control | 21(0/0) | 21(0/0) | 42(0/0) | - | - | TRP | Urine | GC-MS | 5,5 | 5,5 | TRP# |
| 15 | (Gevi, Zolla et al. 2016) | Italy | Autism | Healthy Control | 30(22/8) | 30(22/8) | 60(44/16) | 4.83 (0.30) | 4.83 (0.30) | KA,KYN,QA,XA | Urine | HILIC-UHPLC | 5,5 | 6 | KA*,KYN*,QA#,XA# |
| 16 | (Lim, Essa et al. 2016) | Oman | Autism | Healthy Control | 15(10/5) | 12(10/2) | 27(20/7) | 8.47(2.36) | 9.61(2.9) | KA,KYN,PA,QA | Serum | UHPLC | 7,5 | 6 | KA#,KYN#,PA*,QA# |
| 17 | (Bryn, Verkerk et al. 2017) | Norway | Autism | Healthy Control | 65(52/13) | 30(14/16) | 95(66/29) | 11.2(2.02) | 10.9(2.15) | 3HK,KA,KYN,QA,TRP | Serum | HPLC | 4,5 | 6 | 3HK#,KA*,KYN#,QA*,TRP# |
| 18 | (Bugajska, Berska et al. 2017) | Poland | Autism | Healthy Control | 27(27/0) | 13(13/0) | 40(40/0) | 4.37(2.19) | 5.00(2.74) | Isoleucine,Leucine,Phenylalanine,TRP,Tyrosine,Valine | Plasma | HPLC-UV/VIS | 5,5 | 7 | Isoleucine*,Leucine*,Phenylalanine*,TRP*,Tyrosine*,Valine* |
| 19 | (Li, Shen et al. 2018) | China | ASD | Non-ASD | 27(20/7) | 27(20/7) | 54(40/14) | 5.22(1.09) | 5.56(0.51) | Leucine + Isoleucine,Phenylalanine,TRP,Tyrosine,Valine | Urine | HPLC | 8 | 4,5 | Leucine + Isoleucine*,Phenylalanine*,TRP*,Tyrosine*,Valine* |
| 20 | (Ormstad, Bryn et al. 2018) | Norway | Autism | Healthy Control | 65(52/13) | 30(14/16) | 95(66/29) | 11.2(2.02) | 10.9(2.1) | KA,KYN,QA,TRP | Serum | HPLC | 6,5 | 5 | KA*,KYN#,QA*,TRP# |
| 21 | (Liang, Xiao et al. 2020) | China | Autism | Healthy Control | 22(17/5) | 22(15/7) | 44(32/12) | 7.6(1.8) | 6.4(3.3) | TRP | Urine | HNMR | 5,5 | 5,5 | TRP# |
| 22 | (Olesova, Galba et al. 2020) | Slovakia | Autism | Healthy Control | 24(24/0) | 13(13/0) | 53(53/0) | 4.1(0.8), 7.7(0.9,) | 4.7(0.7), 8.2(1.2) | XA | Urine | LC-MS/MS | 6 | 6 | XA*, XA# |
| 23 | (Bilgiç, Abuşoğlu et al. 2022) | Turkey | Autism | Healthy Control | 68(59/9) | 44(35/9) | 112(94/18) | 35.5m(9.9) | 36.5m(11.8) | 3HAA,3HK,KA,KYN,TRP | Serum | HPLC | 5,5 | 5 | 3HAA*,3HK#,KA#,KYN#,TRP# |
| 24 | (Carpita, Nardi et al. 2022) | Italy | Autism | Healthy Control | 24(17/7) | 24(9/15) | 48(26/22) | 27.75(6.97) | 33.29(8.05) | KA,KYN,QA,TRP | Serum, PRP | ELISA | 5,5 | 9 | KA*,KYN*,QA*,TRP*  KA*,KYN#,QA*,TRP* |
| 25 | (Gagliano, Murgia et al. 2022) | Italy | Autism | Healthy Control | 15(15/0) | 25(16/9) | 40(31/9) | 9(4.28) | 12(2.17) | TRP | Serum | HNMR | 6,5 | 4,5 | TRP* |
| 26 | (Kalejahi, Kheirouri et al. 2022) | Iran | Autism | Healthy Control | 35(24/11) | 31(18/13) | 66(42/24) | 8.10(4.00) | 7.30(2.60) | TRP | Plasma | HPLC | 6,5 | 7 | TRP* |
| 27 | (Raghavan, Anand et al. 2022) | USA | Autism | Neurotypical | 87(68/19) | 326(125/201) | 413(193/220) | - | - | TRP | Serum | LC-MS/MS | 4,5 | 6 | TRP* |
| 28 | (Timperio, Gevi et al. 2022) | Italy | Autism | unaffected siblings | 14(11/3) | 14(11/3) | 28(22/6) | 7.06(0.96) | 6.68(1.28) | KA,KYN | Urine | HPLC | 5,5 | 6 | KA*,KYN* |
| 29 | (Chen, Chen et al. 2023) | China | Autism | Healthy Control | 110(95/15) | 55(45/10) | 165(140/25) | 3.22(1.18) | 3.37(1.23) | TRP | Plasma | LC-MS-MS | 8 | 7 | TRP* |

*: Indicates that patients have reduced level of the measured metabolite compared to healthy control

^#:^ Indicates that patients have increased level of the measured metabolites compared to healthy control

TRP: Tryptophan, KYN: Kynurenine, KA: Kynurenic acid, 3HK: 3-Hydroxykynurenine, AA: Anthranilic acid, 3HA: 3-Hydroxyanthranilic acid, XA: Xanthurenic acid, QA: Quinolinic acid, PA: Picolinic acid, HPLC: High performance liquid chromatography, HPLC-MS/MS: High performance liquid chromatography with tandem mass spectrometry, LC-MS/MS: Liquid chromatography with tandem mass spectrometry, UPLC-MS/MS: Ultra performance liquid chromatography with tandem mass spectrometry, GCMS: Gas chromatography–mass spectrometry, LC: Liquid chromatography, HPLC-UV: High perfomance liquid chromatography- Ultra-violate

**ESF. Table 6**. Results of Meta-regression

| Variables | No. of Studies | Covariates | 1-sided p-value | Z-Value |
| --- | --- | --- | --- | --- |
| TRP/CAAs | 16 | Female Cases | -3.38 | 0.0004 |
|  |  | Sample Size | -1.73 | 0.042 |
| CAAs | 7 | Female Cases |  |  |
| KA/KYN | 4 | Male Cases | 2.18 | 0.014 |
|  |  | Sample size | 2.87 | 0.002 |
|  |  | Fasting (Yes) | -1.77 | 0.038 |
| KYN | 4 | Latitude | -2.62 | 0.0044 |
|  |  | Male Cases | -4.13 | 0.000 |
|  |  | Female Cases | -2.82 | 0.0024 |
|  |  | Sample size | -3.62 | 0.0001 |
|  |  | Fasting-Yes | 5.13 | 0.0000 |
| KA | 4 | Female-Case | -2.23 | 0.0130 |

TRP: Tryptophan, KYN: Kynurenine, KA: Kynurenic acid, CAAs: Competing amino acids.

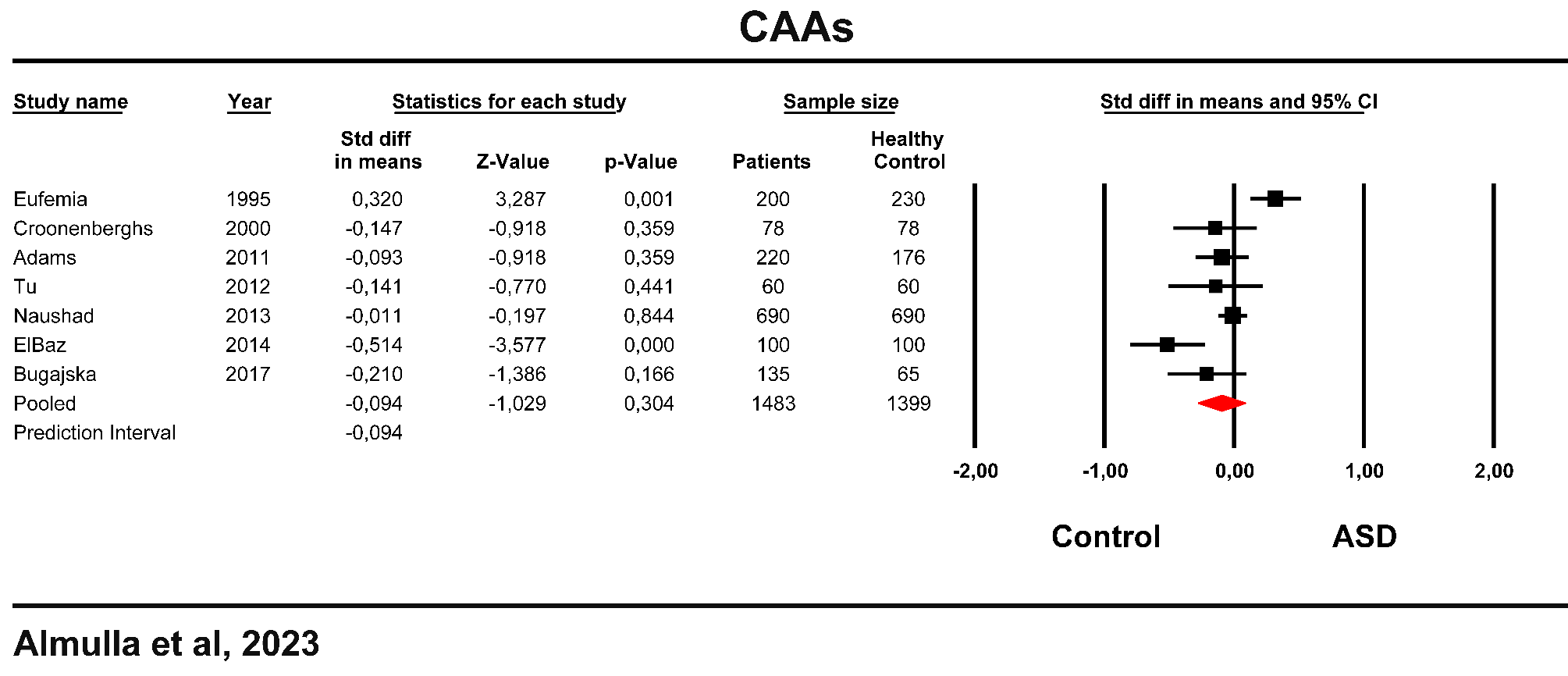

**ESF, Figure 1**. Forest plot of competing amino acids (CAAs) in the patients with autism spectrum disorder (ASD) versus healthy control.

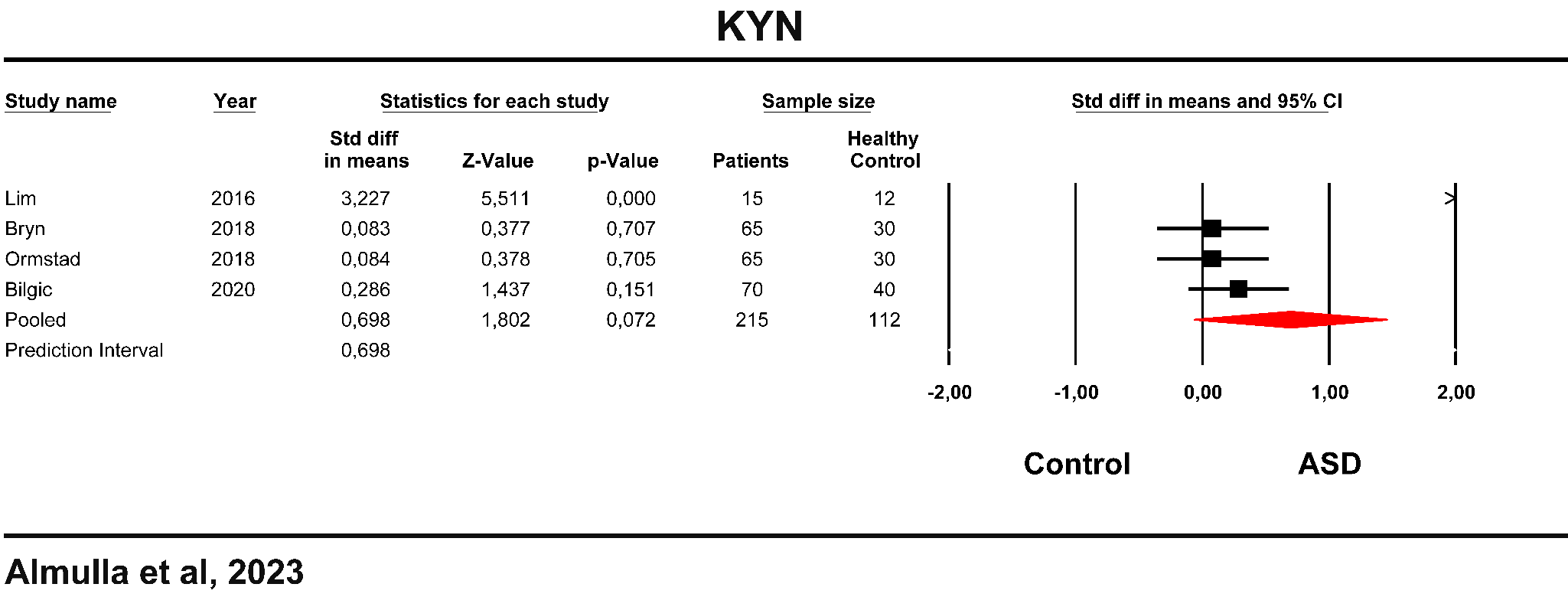

**ESF, Figure 2**. Forest plot of kynurenine in the patients with autism spectrum disorder (ASD) versus healthy control.

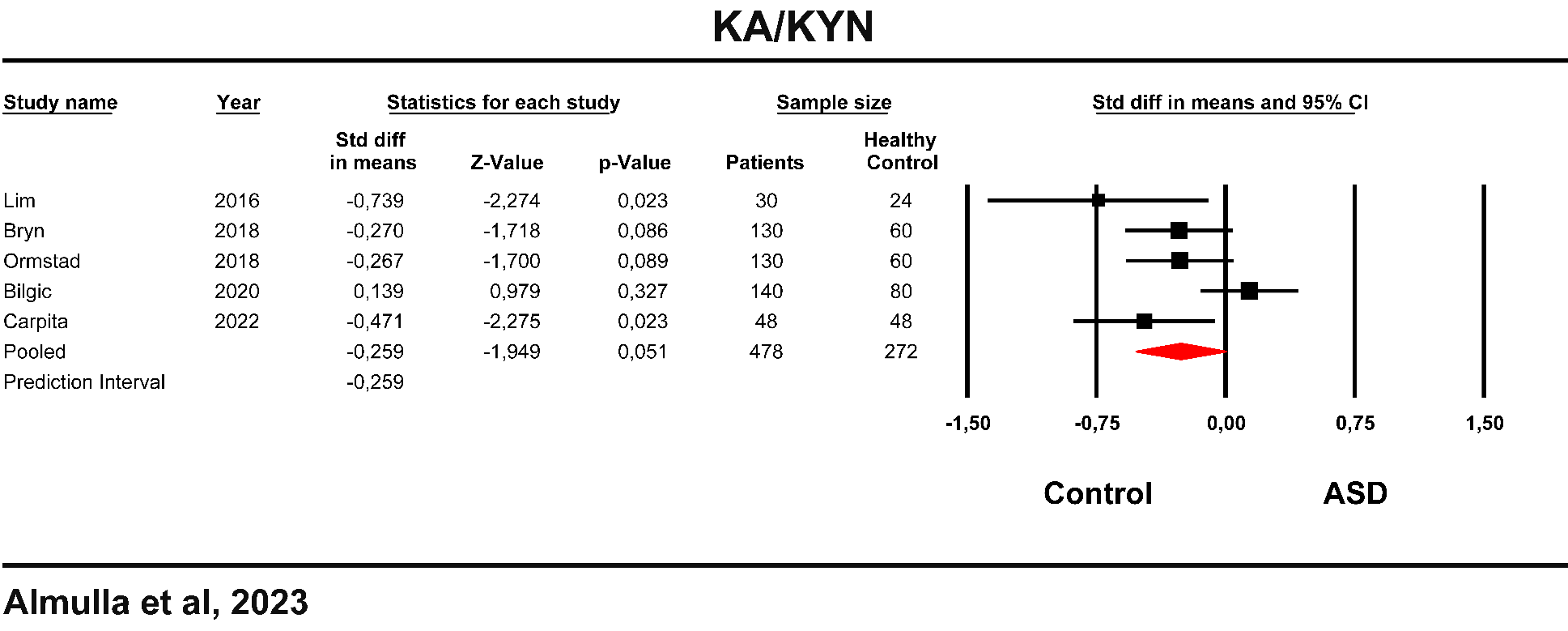

**ESF, Figure 3**. Forest plot of kynurenic acid (KA)/Kynurenine (KYN) in the patients with autism spectrum disorder (ASD) versus healthy control.

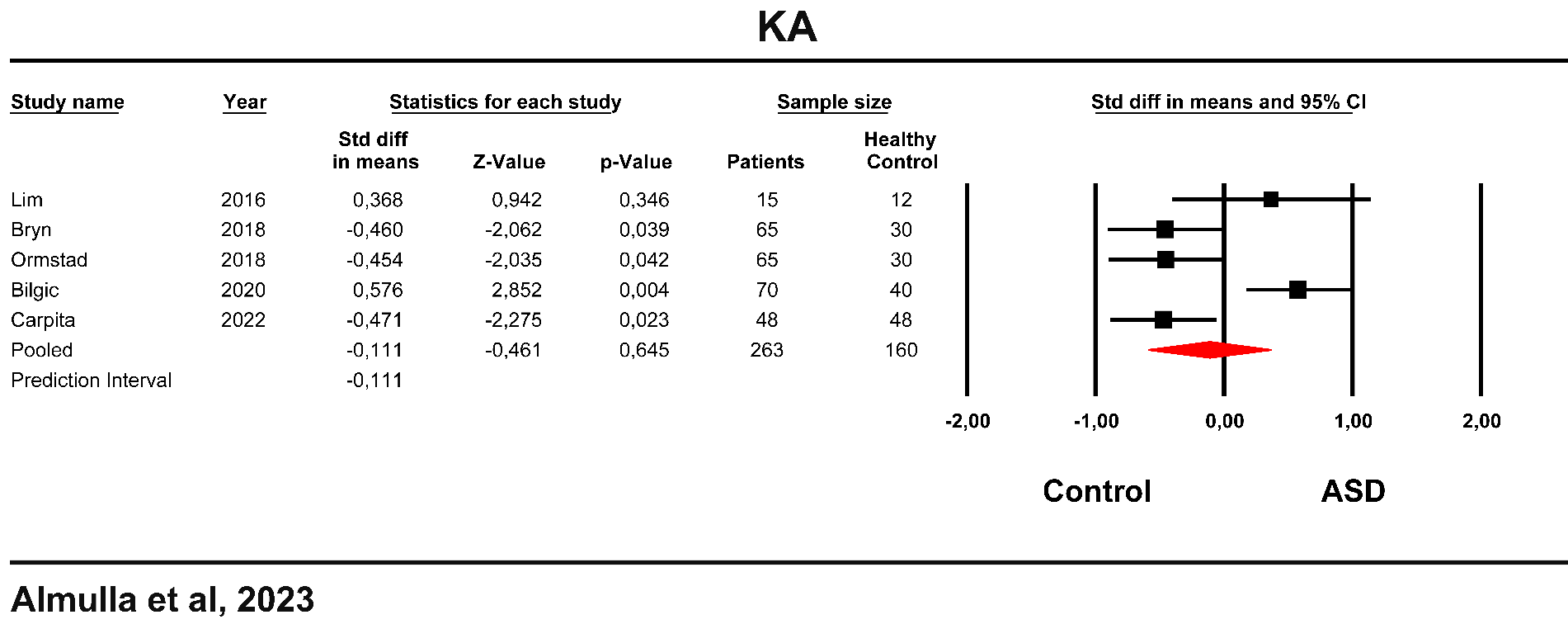

**ESF, Figure 4**. Forest plot of kynurenic acid (KA) in the patients with autism spectrum disorder (ASD) versus healthy control.

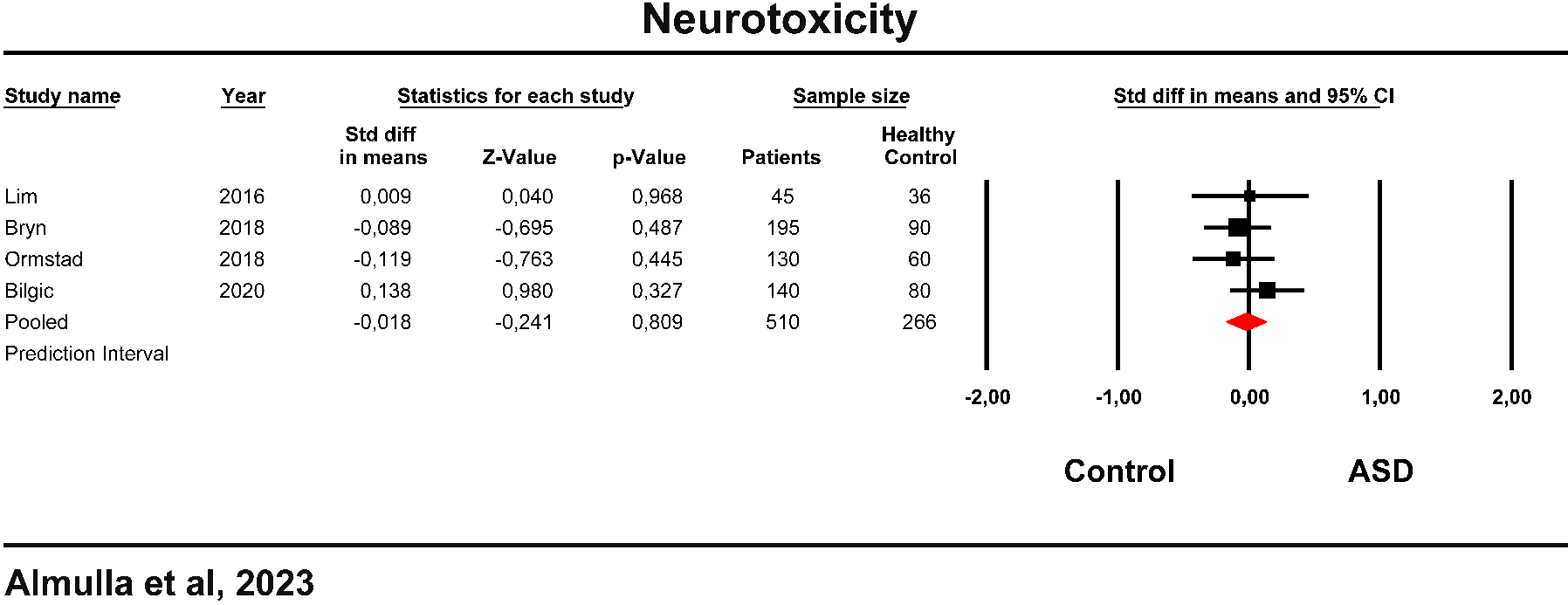

**ESF, Figure 4**. Forest plot of neurotoxicity (KYN+3HK+QA+XA+PA) in the patients with autism spectrum disorder (ASD) versus healthy control.

**Abbreviation:**

TRP: Tryptophan

KYN: Kynurenine

KA: Kynurenic acid

3HK: 3-Hydroxykynurenine

AA: Anthranilic acid

3HA: 3-Hydroxyanthranilic acid

XA: Xanthurenic acid

QA: Quinolinic acid

PA: Picolinic acid

CAAs: Competing amino acids

IDO: Indoleamine 2,3 dioxygenase

TDO: Tryptophan 2,3 dioxygenase

KAT: Kynurenine aminotransferase

KMO: Kynurenine 3-monooxygenase

TRYCATs: Tryptophan Catabolites

TRYCAT pathway: Tryptophan catabolite pathway

SMD: Standardized mean difference

IL-6: Interleukin-6

IFN-γ: Interferon-gamma

O&NS: Oxidative and nitrosative stress

MOOSE: Meta-Analyses of Observational Studies in Epidemiology

CSF: Cerebrospinal fluid

SD: Standard definition

IOR: Interquartile range

ICS: Immune confounder scale

CI: Confidence intervals

LC-MS/MS: Liquid chromatography with two mass spectrometry

HNMR: Proton nuclear magnetic resonance

HPLC: High-performance Liquid-Chromatography.

NMDA: N-methyl-D-aspartate

ELISA: Enzyme-Linked immunosorbent Assay
